# Health service improvement using positive patient feedback: systematic review and change model

**DOI:** 10.1101/2022.09.10.22279800

**Authors:** Rebecca Lloyd, James Munro, Kerry Evans, Amy Gaskin-Williams, Ada Hui, Mark Pearson, Mike Slade, Yasuhiro Kotera, Giskin Day, Joanne Loughlin-Ridley, Clare Enston, Stefan Rennick-Egglestone

## Abstract

**Background:** Patients, families, and communities regularly provide feedback about care and treatment received from healthcare services, most of which is positive. The aim of this review was to examine how positive feedback creates change within healthcare settings.

**Methods:** Included documents were empirical studies where the full text is publicly available in English, and where a change has occurred within healthcare services attributed to positive feedback from service users, their families, or the community. They were identified through database searches (ACM Digital Library, AMED, ASSIA, CINAHL, MEDLINE and PsycINFO), forwards and backwards citation, and expert consultation. Data was synthesised into a change model describing forms, mediators and moderators of change. A protocol was prospectively registered: https://osf.io/5x46c.

**Results:** Sixty-eight papers were included, describing research conducted in 32 countries across six continents, with qualitative (n=51), quantitative (n=10), and mixed (n=7) methods. Only two described interventional studies. The most common form of feedback was ‘appreciation’ (n=28). The most common recipients were nurses (n=29). Positive feedback was most commonly given in hospitals (n=27) and community healthcare (n=19). Positive feedback mostly led to positive outcomes categorised as (a) short-term emotional change for healthcare workers (including feeling motivated and improved psychological wellbeing), (b) work-home interactional change for healthcare workers (such as improved home-life relationships), and (c) work-related change for healthcare workers (such as improved performance and staff retention). Undesirable changes included embarrassment when receiving feedback, tension in the patient-professional relationship, and envy when not receiving positive feedback.

**Conclusion:** Positive feedback can provide the opportunity to create meaningful health service change. Healthcare managers may wish to use positive feedback more regularly, and to identify and address barriers to staff receiving feedback. Further interventional research is required to establish the effectiveness of receiving positive feedback in creating change, and to understand the influence of feedback content.

## Introduction

The term ‘feedback’ often refers to the process whereby the effect of an action is fed back to change future actions. Feedback can be positive, where the gap between actual and desired experience is narrow, for example, when healthcare treatment aligns with patient wishes. Feedback can also be negative, where the gap between actual and desired experience is wide (1). For example, when healthcare treatment falls short of patient wishes. People working in health services regularly receive feedback about the treatment provided to patients, including from the patients themselves, family members, and informal carers (2). Patient feedback differs from patient engagement, which can be defined as patients taking an active role in their healthcare experience to meet a particular objective, such as accessing additional support groups (3).

The most frequent form of feedback from healthcare service users is informal feedback, usually exchanged through conversations day-to-day (2), but also via letters and online forums (4). Formal feedback is a structured evaluation, often collected through methods such as national or local surveys (5). Both types of feedback can be given in-person or online and can include generic content (such as ‘fantastic care’), or specific (such as ‘the food was fantastic during my hospital stay’) (6). Some feedback is solicited, such as staff requesting patients to complete feedback surveys (7), whereas some is unsolicited (8).

Positive feedback is much more common than negative feedback, with a linguistic analysis of comments posted on the UK’s National Health Service (NHS) Choices website finding positive evaluations to be three times as likely as negative (6). Positive feedback tends to be shorter, often expressed just as a single word such as ‘fantastic’ (9), and may be conceptualised as including material displays, such as gift-giving, cards, and donations to healthcare services (10). Positive feedback is evident in a variety of forms, such as favourable responses to surveys (4), online comments (9), compliment letters (11), and informal thanks (4). Positive and negative evaluations may also be given in combination, forming ‘mixed’ feedback (12). Increasingly, feedback is received through online sources, with online feedback being mostly positive in tone (13). Expressions of positive feedback may differ culturally; British culture tends to be less emotionally expressive than American culture (14). Eastern cultures tend to value low arousal emotions such as peacefulness, whereas Western cultures tend to value high arousal emotions such as happiness (15).

One purpose of collecting service user feedback is to assist with performance monitoring and assurance, such as comparison between healthcare providers, impact of service changes, informing commissioning decisions, and compliance with standards. Another purpose is to share understanding and information to assist with service choices for other service users, highlight service problems and hold services accountable, and increase staff understanding of service user experience (16).

Service users may want to give positive feedback to acknowledge, reward, and promote desired behaviour in healthcare staff (11). This frequently includes expressions of gratitude, conceptualised as the communication of an emotion or state which signals recognition that others have done something to benefit us (17). Expressions of gratitude and positive feedback are interrelated, but not synonymous. Gratitude can describe an individual’s attitude, moral values, daily habits, or emotional state, with the intention of reciprocity often at its core (17).

In some cases, expressions of gratitude can serve as a positive evaluation of an individual or group accomplishment, and therefore be a form of positive feedback. For example, grateful postcards and letters sent to palliative care units from patients and families recognised the care and treatment received, the value of palliative care, and offered messages of support and encouragement about the service (18). Similarly, throughout the COVID-19 pandemic, many healthcare service users used Twitter to express their gratitude for the work, effort, saving and caring of healthcare staff and services (19). However, not all expressions of gratitude will be given with the intention of recognising accomplishments, such as habitually thanking healthcare staff in the expectation that it would ensure continuation of good treatment (20). Similarly, not all positive feedback will include expressions of gratitude, with some offering objective descriptions of excellent care and treatment practices. The current review will characterise expressions of gratitude towards healthcare staff as a type of positive feedback, acknowledging how these concepts interrelate and discriminating between them where possible.

Patient feedback is given in abundance and the use of readily available patient feedback may offer a cost-effective way to create meaningful change within healthcare services (4). In one case study, service user accounts of distress during admission to mental health inpatient services were used as a resource to inform service improvement work. An 80% drop in complaints was observed over the following 14 months after implementation (21). Similarly, the NHS in England uses the Friends and Family Test (FFT) to collect information about patient experience of what is going well and what needs improving (22). The FFT was able to highlight positive patient views about remote appointments during the pandemic. In Buckinghamshire Healthcare NHS Trust, for example, patients felt that video or telephone appointments were safer, saved time, removed need for finding and paying for parking, allowed more time for talking, was better for the environment, and were more likely to be on time (23). This provides valuable insight in how patients are experiencing changes to healthcare services. In Japan, healthcare workers also reported that positive communication and acknowledgement, including from patients, acted as a mental health resource during the Covid-19 pandemic (24).

Formal processes exist within healthcare services, such as the UK’s NHS, to use service user feedback. In the UK, registered healthcare professionals are often required to collect and reflect on service user feedback as a formal requirement of practice. The UK Nursing and Midwifery Council (NMC) describe the process of revalidation required for all nurses and midwives in the UK to maintain their registration with the NMC, promote good practice, and strengthen public confidence (25). The revalidation process requires five pieces of feedback which can come from a variety of sources and forms, including from service users. Similarly, the General Medical Council (GMC) require reflection on feedback from service users at least once in each revalidation cycle, which is required every five years (26). Doctors with the GMC are encouraged to reflect on feedback from existing sources of patient feedback, such as letters, cards, or team feedback, and should cover the whole scope of their practice. However, the Health and Care Professionals Council (HCPC) standard guidelines state that asking for, receiving, and reflecting on service user feedback is not a specific requirement, but can be useful for continuing professional development (CPD) (27). This feedback refers to evidence that changes made in line with CPD standards have been beneficial to the service user, such as letters produced for a CPD audit. Positive feedback from service users therefore has the potential to be used in current formal practice requirements for UK healthcare professionals. However, some have described the process as a ‘hoop jumping’ exercise with no impact on patient care, and some take issue with using feedback because patient-facing working patterns do not create equitable patient-facing opportunities (28).. The infrequency at which patient feedback is required for formal processes may contribute to the view that patient feedback is unrepresentative and tokenistic. Despite expressing sceptical views, healthcare staff have described its value as beneficial (28).

Feedback can also be used for quality improvement. Aggregating feedback can provide insight into quality of care for service users and highlight real-time priorities for service inspections and improvement (29). The NHS promotes two models for improving quality by implementing changes (30). The first is the five-step improvement approach, which offers a systematic framework for improvement projects. The five stages are preparation (preliminary planning such as defining aims, objectives, and collecting baseline data), launch (the official start of a project), diagnosis (understanding the current process and defining the problem being addressed), implementation (testing potential solutions to the problem), and evaluation (achievements and learning are captured to make improvements the norm in that service). The second quality improvement model fits into the implementation phase of the five-stage improvement approach. The model for improvement asks three preliminary questions: what are we trying to accomplish? How will we know that a change is an improvement? And what changes can we make that will result in the improvements that we seek? The model consists of four components: plan, do, study, act (PDSA). Improvements can be made by planning how to test change, carrying out what has been planned, measuring the outcomes of the test, and acting on the results to modify and improve services (30).

However, there are a range of organisational barriers to the effective use of patient feedback by health services (31). Staff can lack the time or skills required to interpret formal feedback (4), and might be reluctant to engage with feedback communicated informally through online platforms such as Facebook or Twitter (9, 32). In some contexts, online feedback is emerging at a faster rate than health services could respond to (16). An example is Care Opinion, an online service for the collection of feedback that enables staff responses, but conversations are often closed with a ‘thank you’ in response to positive feedback (4) rather than with an account of how this feedback was used to create change. Even where informal feedback is acted on by healthcare staff, the improvements made are often informally implemented in real-time and hence are not captured by quality improvement methods (33). Formal methods of processing service user feedback are also focussed on complaints in healthcare and positive feedback is often overlooked as a useful source of feedback (34). Healthcare staff may assume that feedback is negative in tone (13), and can dismiss or fail to value positive feedback (9). The lack of credibility afforded to positive feedback may be due to its tendency to be shorter and may be viewed by staff working in quality improvement as lacking direct measurability compared with clinical outcomes or complaints (33).

Three reviews have investigated the value of gratitude in healthcare settings (35-37). A meta-narrative review of 56 studies investigated gratitude in healthcare with a particular focus of interpersonal experiences (36). The review described how gratitude can act as ‘social capital’ as it empowers and motivates recipients through strengthened social bonds, connectedness, and an increased willingness to reciprocate. Day (2020) also highlights how patient gratitude can benefit staff wellbeing, such as being protective against burnout and having physical health benefits and may be an indicator of quality of care. A scoping review (35) included 32 studies from three databases, and examined the characteristics, focus, and effects of gratitude. It found that gratitude influenced healthcare professionals professionally and personally, generating positive feelings such as pride, satisfaction, and a sense of wellbeing. It also generated reciprocal gratitude among other healthcare professionals. The review highlighted a limited evidence base and concluded that a systematic investigation into the effects of patient gratitude was needed, in line with Medical Research Council (MRC) recommendations on intervention development processes (38).

A narrower systematised review on the impact of gratitude in healthcare settings included 23 studies from three databases (37). The review found one harmful change, where service user gift-giving resulted in healthcare staff feeling tension and pressure to meet patient expectations, undermining the service user-professional relationship. The review found that patient gratitude can also create helpful changes for healthcare staff, identified as work-related change (such as improved team performance and work-related satisfaction), direct benefits to staff health (such as increased sleep quality and decreased headaches), and proximal emotional change (such as feeling rewarded, proud, motivated, and fulfilled). In some cases, change was mediated by team information sharing, and was moderated by the psychological demands of the job role. A preliminary change model was produced as a result of the review findings which presented a summarised version of the change process. Following expressions of gratitude from patients, healthcare staff experience a change in their perceptions, either within their relationships with patients or colleagues or the way they experience work. Psychological demands of staff job role moderated this change. A change in perception lead to the harmful and helpful changes previously described, with helpful changes mediated by work-related satisfaction and increased information sharing.

### Aims and objectives

Whilst three reviews have examined patient gratitude in healthcare settings, none has examined the impact of positive feedback more broadly. The aim of this review is to examine how positive feedback received by health services about care and treatment can create change within healthcare settings. The objectives were (1) to identify measures used to quantify change; (2) to create a model describing types of change and how it occurs; and (3) to identify recommendations for the collection and use of positive feedback by health services.

## Methods

A systematic review of empirical studies was conducted. Positive feedback was defined as a response from healthcare service users, families or the community indicating concordance between desired and actual experiences regarding their care or treatment, delivered to healthcare staff or systems. The 2021 update of the Preferred Reporting Items for Systematic Reviews and Meta-Analyses (PRISMA) checklist was used in reporting the review (39). A review protocol was prospectively registered with the Open Science Framework (https://osf.io/5x46c), which extended the protocol for a prior review (37), with extensions validated through a scoping review. A narrative synthesis of review data was performed to create a preliminary model of change (40).

### Search strategy

#### Electronic database searches

Databases were selected to cover a range of domains relating to healthcare service delivery. Searches were conducted from inception to 18^th^ March 2022 on PsycINFO, AMED, MEDLINE, CINAHL, and the ACM Digital Library (ACM DL), and from inception to 15^th^ December 2021 on ASSIA (the shorter date was due to a constraint in institutional access). The ACM DL indexes papers where computation and human interaction with technology is a primary focus and was included as feedback is frequently collected via electronic systems.

Search terminology was extensively tested during a previously conducted systematised review focusing on expressions of patient gratitude (37), extended for the current review to encompass positive feedback beyond gratitude and healthcare systems more generally, and informed by the learning from the scoping searches. Scoping searches identified terms which were synonymous with ‘positive feedback’, such as ‘positive evaluation’ and ‘praise’, and terms which described healthcare systems, such as ‘healthcare services’ and ‘healthcare communities’.

Search terms which linked less closely to positive feedback but produced a high volume of documents, such as recognition, were searched in titles only. In the initial filter by title, the screening team took care not to exclude papers in the event of ambiguity.

The following search strategy was used for MEDLINE, PsycINFO, and AMED (all searched through Ovid):

1. Health* staff.ti,ab.
2. Health* worker*.ti,ab.
3. Medical staff.ti,ab.
4. Medical worker*.ti,ab.
5. Exp Health Personnel/
6. Health* system*.ti,ab.
7. Health* service*.ti,ab.
8. Health* organi#ation*.ti,ab.
9. Health* communit*.ti,ab.
10. 1 or 2 or 3 or 4 or 5 or 6 or 7 or 8 or 9
11. Grat*.ti,ab.
12. Appreciat*.ti,ab.
13. Recog*.ti.
14. Thank*.ti.
15. Positive* feedback.ti,ab.
16. Positive* evaluat*.ti,ab.
17. Praise*.ti,ab.
18. 11 or 12 or 13 or 14 or 15 or 16 or 17
19. 10 and 18
20. Remove duplicates from 19

This search strategy was amended for CINAHL and ASSIA (amendments in S1 File).

The ACM Digital Library only allows searches constructed using combinations of keywords, which generates a series of online pages of possible matches in order of relevance. Keyword combinations were identified from the MEDLINE search strategy (searches in S1 File). For each keyword combination, results pages were sequentially inspected for potentially includable documents, and inspection was discontinued when three subsequent pages of non-relevant results were observed.

When developing the search strategy, documents from the prior review (37) were used as marker papers to evaluate search strategy sensitivity.

#### Citation tracking

Reference lists for included documents were manually inspected for further includable documents (backwards referencing). Forward referencing of included documents was conducted using Google Scholar. Forward and backward citation was repeated on additional included documents until no further documents were included.

#### Expert consultation

Once the final list of includable documents from electronic databases was identified, three experts in healthcare service delivery were asked to identify any potentially includable documents which had been omitted. Experts consisted of a healthcare manager responsible for feedback, an academic expert, and a technology creator who collects feedback about healthcare. Proposed documents were inspected for inclusion by the researcher. Forwards and backward referencing was conducted on additional included documents identified during expert consultation and repeated until no further documents were included.

### Document inclusion

The Population, Intervention, Comparison, Outcome, Study Design (PICOS) search tool was used to specify inclusion (41).

#### Study design

Included documents were empirical studies where the full text is publicly available in English, with a clearly defined research method. Documents were included where a change has occurred within healthcare services attributed to positive feedback from service users, their families, or the community. Change was inclusive of individual healthcare staff changes, such as behavioural, emotional, and attitudinal shift, and systematic or procedural change within healthcare structure.

Documents describing systematic, literature, or scoping reviews, policy statements, conference abstracts, protocols, and documents presented in a blog format were excluded. Documents were excluded where it was unclear whether change occurred as a result of positive feedback, where the identified change preceded positive feedback or directionality was ambiguous (e.g., where a change in healthcare staff or systems caused positive service user feedback), or where the impact of positive feedback was not presented as a study finding but was briefly mentioned as a discussion point.

#### Context

Included documents described research in the context of a healthcare setting, defined as any formal service where healthcare is being delivered, such as in hospitals, outpatient services, hospices, healthcare education, or correctional medical facilities. This was not limited to private or public healthcare services. Documents describing community healthcare settings were also included if staff were providing a formal healthcare service in the community. Documents were excluded where they describe positive feedback occurring within a healthcare system in relation to research being conducted, such as feedback about participation in a randomized clinical trial.

#### Intervention

Included documents described the voluntary expression of positive feedback from healthcare service users, their families, or community members, relating to the care or treatment provided, with healthcare workers or healthcare services as recipients. This included positive feedback expressed verbally and in invariant forms (such as in writing), and positive feedback provided both in-person and remotely (such as online). Expressions of gratitude were included as they may indicate service user feelings about care and treatment and hence can be used as a source of information by healthcare staff or systems. Studies describing ‘recognition’ of healthcare staff or services in relation to appreciation of care and treatment provided were included.

Documents were excluded if (1) the type of service user feedback was not identified as positive, was negative or mixed, ambiguous, or was hypothetical (2) the source of positive feedback was not healthcare service users, families, communities, or was ambiguous (3) positive feedback from healthcare service users, families, or communities was not distinct from feedback provided by peers or the organisation, or (4) expressions of positive feedback were not voluntary (for example, where service users felt that their care and treatment may be negatively impacted if they do not express positive feedback). Feedback was assumed to be given voluntarily unless otherwise stated. Documents describing recognition awards or honours informed by the treatment and care experiences of healthcare service users, such as the Diseases Attacking the Immune System (DAISY) Award (42), were excluded. Similarly, documents describing feedback given via Appreciative Inquiry (a strength-based approach to creating change with a focus on appreciation and positive conversations) were excluded if service user involvement was not explicitly stated or distinguishable from peer or organizational feedback (43). Documents describing donations or gifts to healthcare services were excluded if the motivation for donation was not explicitly described as positive feedback or gratitude towards the healthcare staff or system (37). Studies which describe positive recognition of healthcare staff regarding social status rather than care or treatment provided, such as community support, approval, acceptance, or respect, were excluded (44). Studies were also excluded where healthcare service user satisfaction with care and treatment was described, but not explicitly delivered as positive feedback to healthcare staff or services.

#### Participants

Included documents described participants as working within a formal healthcare environment. The following were in scope: paid or volunteer workers within any healthcare system worldwide; students carrying out a formal healthcare role as part of their studies. Documents describing research into healthcare systems at an organizational level (e.g., where there were no staff participants) were also included. Healthcare systems were defined as any healthcare structure delivering care services to healthcare users.

Documents were excluded where authors did not state whether feedback was provided within a healthcare setting, if participant roles were informal such as unpaid familial caregivers, or if participants were unable to receive feedback.

### Analysis plan

Documents from database searches were exported to EndNote (45) and duplicates were removed. Documents were screened for eligibility, filtered on title in stage one and abstract in stage two. Concordance checking was conducted on a randomly selected 20% of exclusions by a second researcher [SRE] for both stages (title and abstract) of exclusion. Selection processes were piloted until a concordance rate of 95% was achieved on exclusions. Stage 3 screened remaining documents for eligibility based on full text. Retrieved documents were reviewed for inclusion by two researchers, with 100% concordance required on inclusions and exclusions for Stage 3. Uncertainty about the eligibility of a document from both researchers led to it being carried forward to the next stage of screening. At Stage 3, reasons for exclusion were recorded and agreement was required between RL and SRE.

#### Data abstraction and synthesis

A data abstraction table (DAT) was modified from the systematised review (37). The DAT was amended for the purpose of the current systematic review and was piloted using a small number of includable documents to ensure appropriate and efficient design.

The DAT categorised information about study context (such as country of study and healthcare setting), study methodology (such as measures and purpose of measures), recipients and givers of feedback (including healthcare role and healthcare service), type of positive feedback considered, and forms of observed change. For types of feedback, donations were recorded under the higher category of ‘gifts’. The observed change was split into sections: outcomes, mechanisms, moderators, facilitators, barriers, and mediators.

Outcomes were defined as observed changes that have occurred following positive feedback. Mechanisms were defined as processes which produce change. Moderators were defined as factors which alter the degree of change following positive feedback. Facilitators were defined as factors enhancing the observed change. Barriers were defined as factors impeding the observed change. Mediators were defined as factors creating an indirect pathway between two variables for the change process to occur (46).

Specific links between outcomes, mechanisms, mediators, moderators, facilitators, and barriers were retained in the DAT, for example if an included document presented evidence that a specific outcome was produced by a specific mechanism. Items were listed in all relevant categories where there was variation in categorisation among studies. Where papers reported more than one study within a single paper, only data from relevant studies were extracted. Quality assessment score was also reported in the DAT, and if a section of the DAT was not clearly stated in a document, it was recorded as ‘N/A’.

#### Change model development

Each category was assessed for similar items and were combined to develop a synthesised typology. A preliminary change model was produced to describe outcomes, mechanisms, mediators, and factors enhancing or supporting change. Moderators, facilitators, and barriers were combined into two tables reflecting factors that enhance change and factors that hinder change. The change model was reviewed by an expert panel consisting of national and local health service representatives experienced with working with feedback, the director of a company providing a public online feedback platform (JM), and three experienced researchers.

#### Subgroup analysis

Subgroup analyses were planned for documents which identify change through expressions of healthcare service user gratitude, and for documents which were assessed as having high quality methodology. Quality of included documents was assessed using the Mixed Methods Appraisal Tool (MMAT) due to its rigorous assessment of mixed methods study methodologies and high applicability (47). An additional unplanned subgroup-analysis compared mainly public versus mainly private healthcare settings.

## Results

### Summary of papers

Database searches identified 17,619 records once duplicates were removed. Sixty-eight papers were included (see Fig 1). The PRISMA checklist is in S2 File.

**Fig 1.**
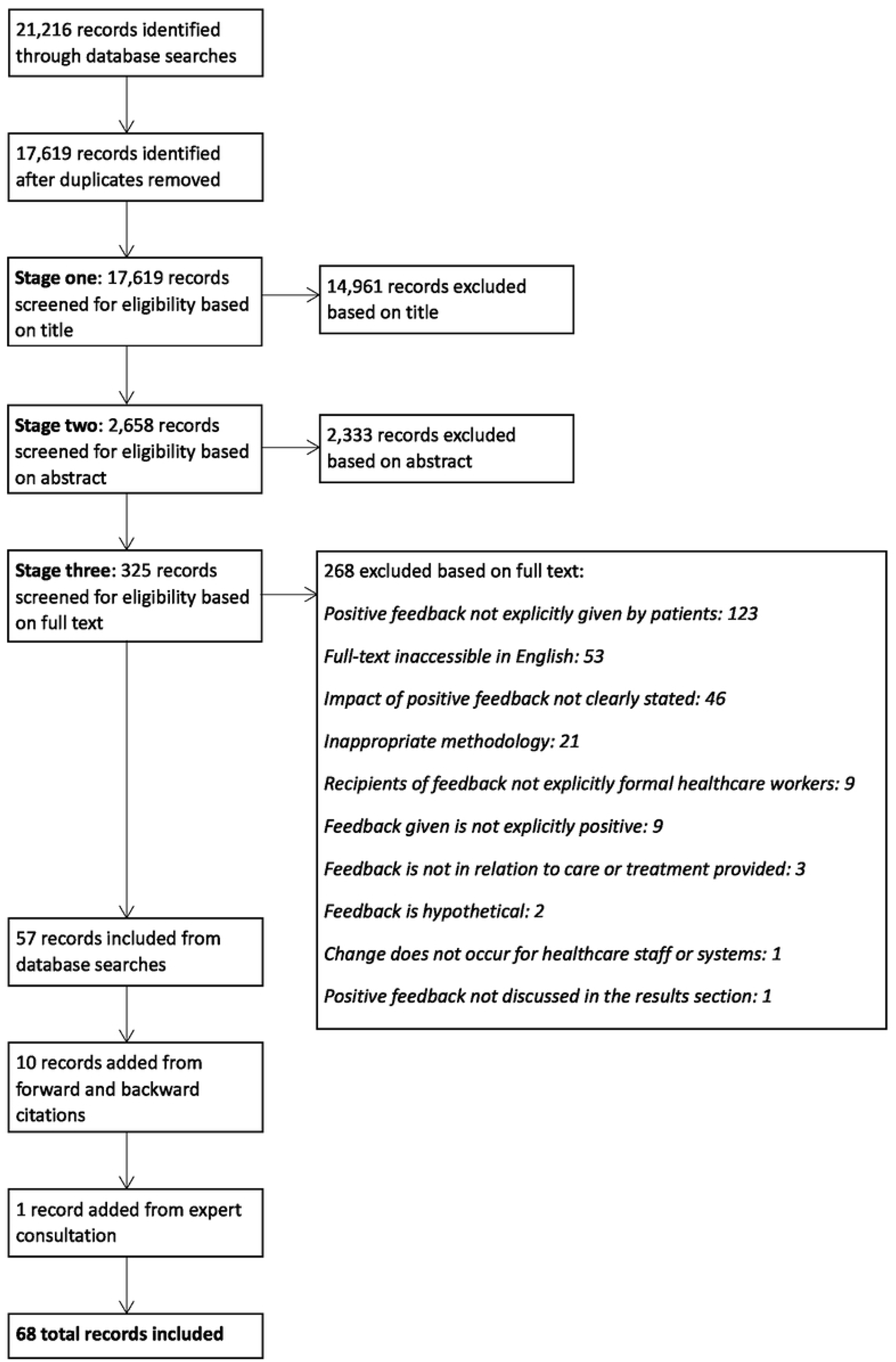
PRISMA Flow Diagram of Included and Excluded Screening Process.

### Characteristics of included studies

A summary DAT is in Table 1, and the full DAT is in S3 Table. One included study presented methodology and results across two papers (48, 49) which were merged to form one record [ID 67]. Where several papers were created from a single study, these were considered companion papers. Three studies had corresponding companion papers [ID 3 and 4; ID 11 and 12; ID 45 and 46].

**Table 1.**
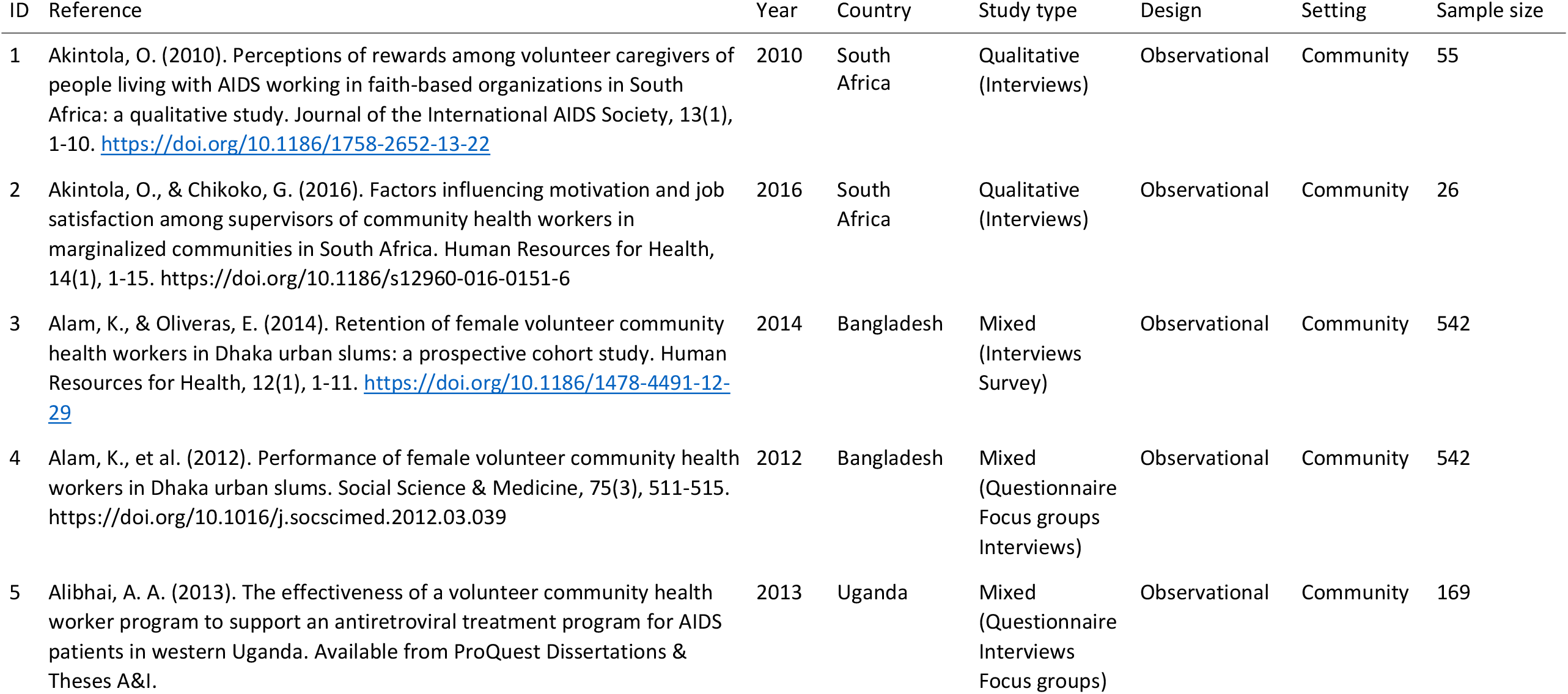
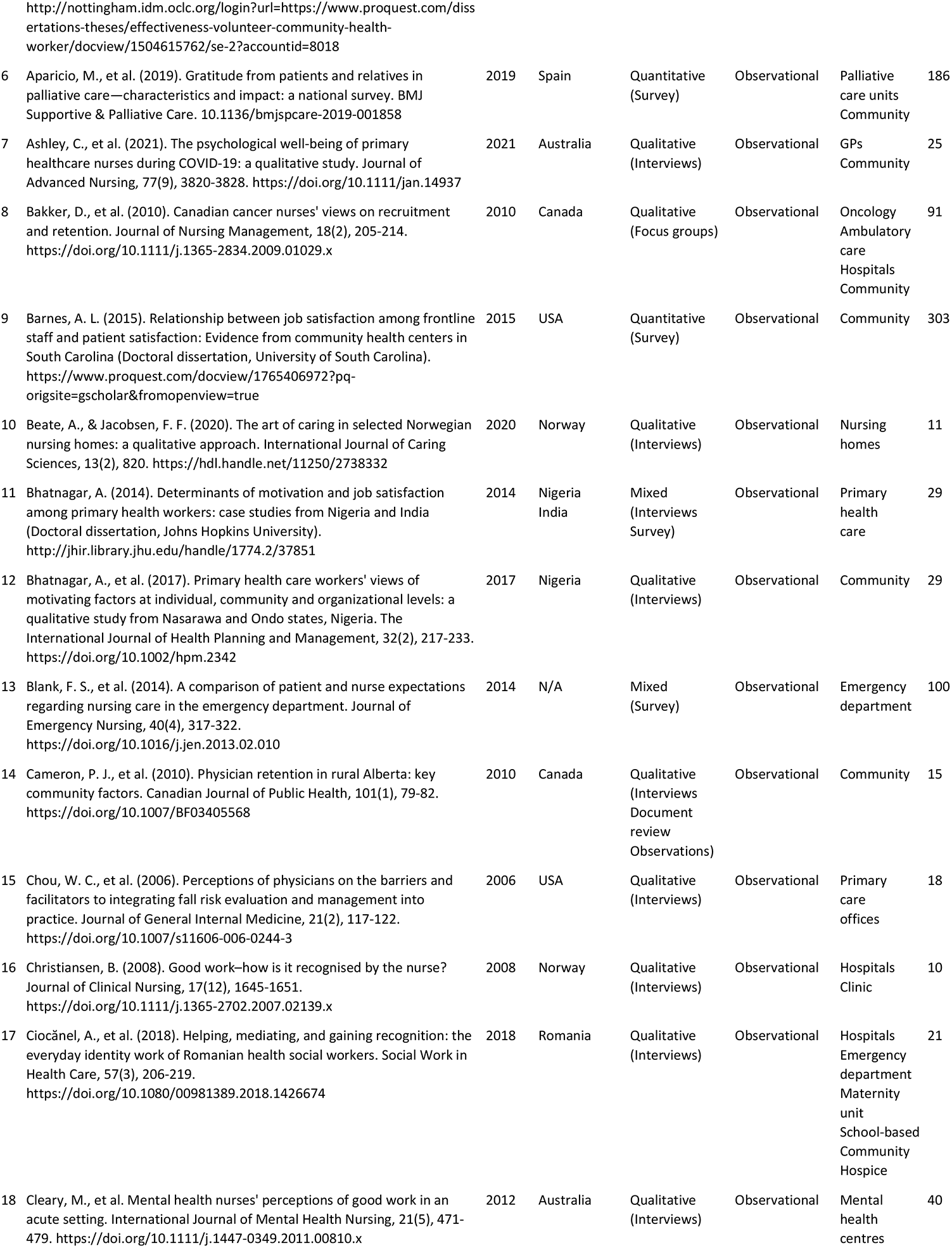
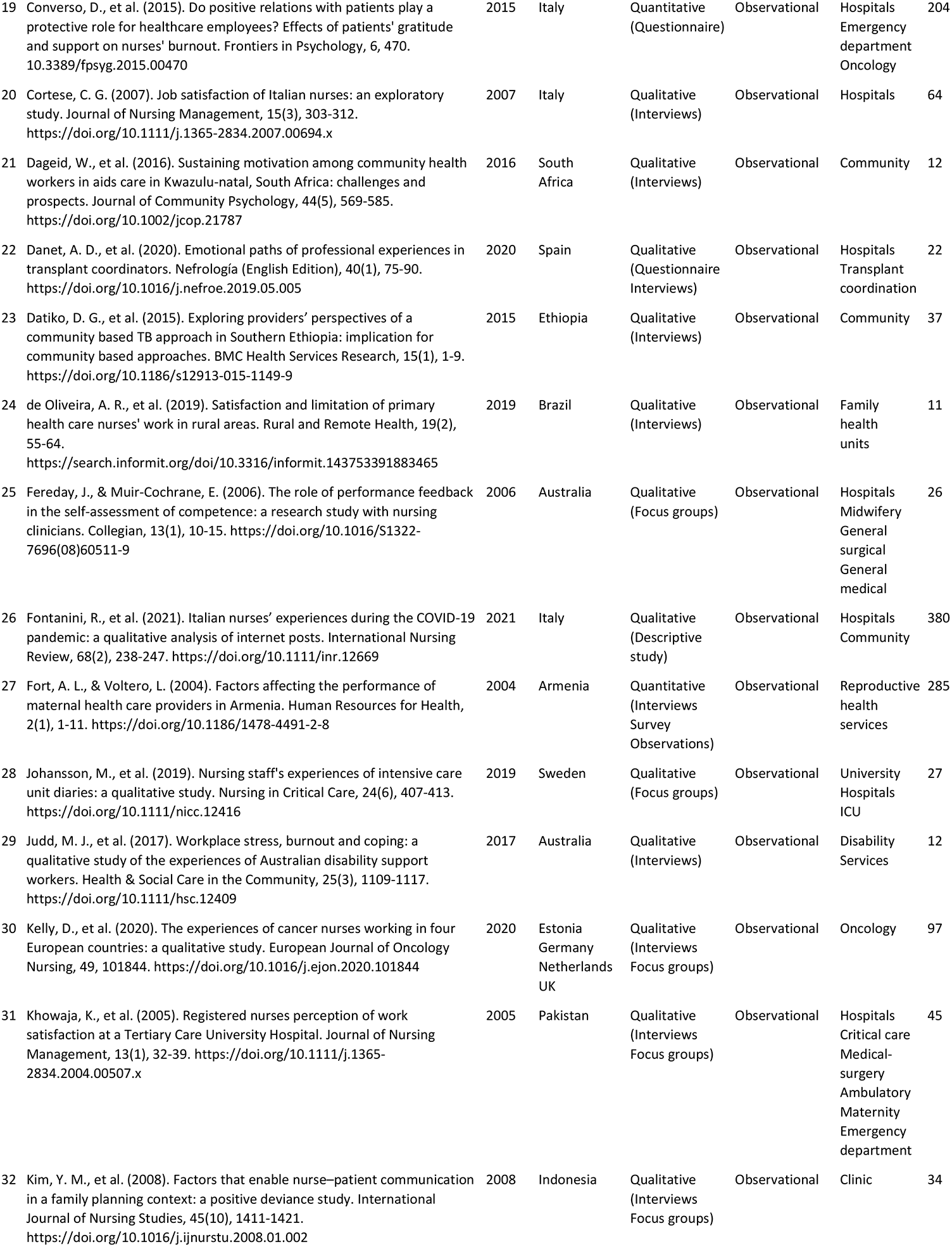
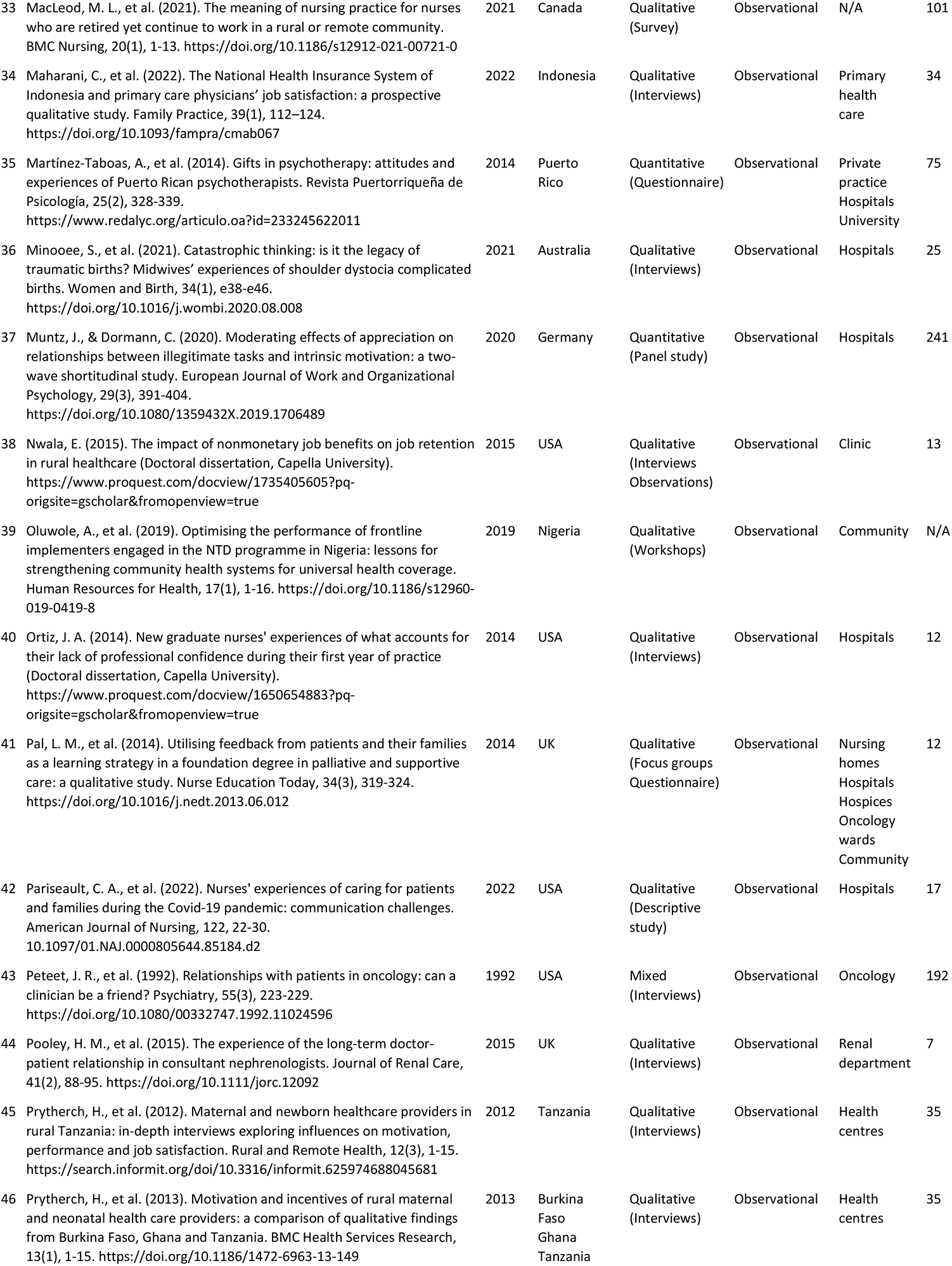
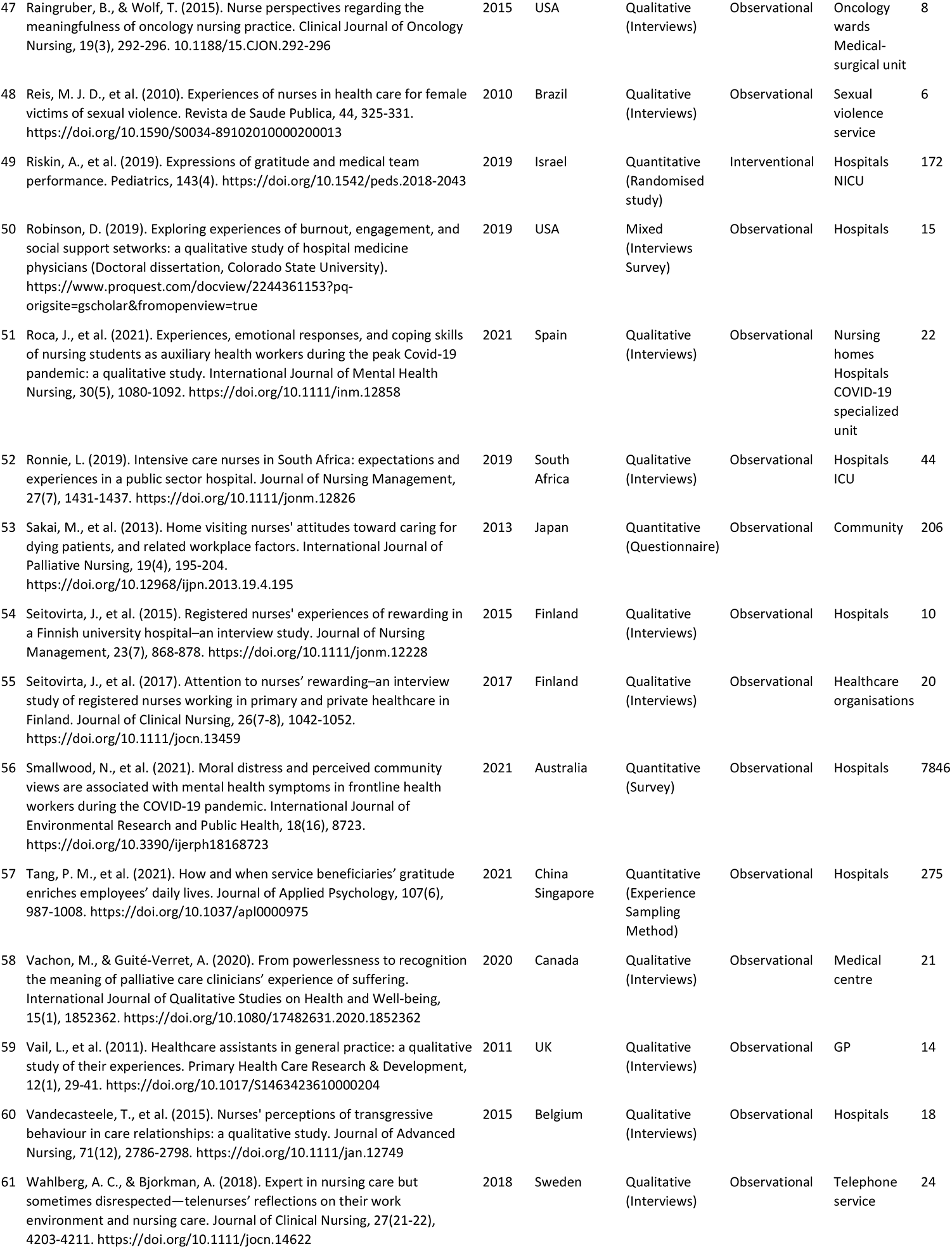
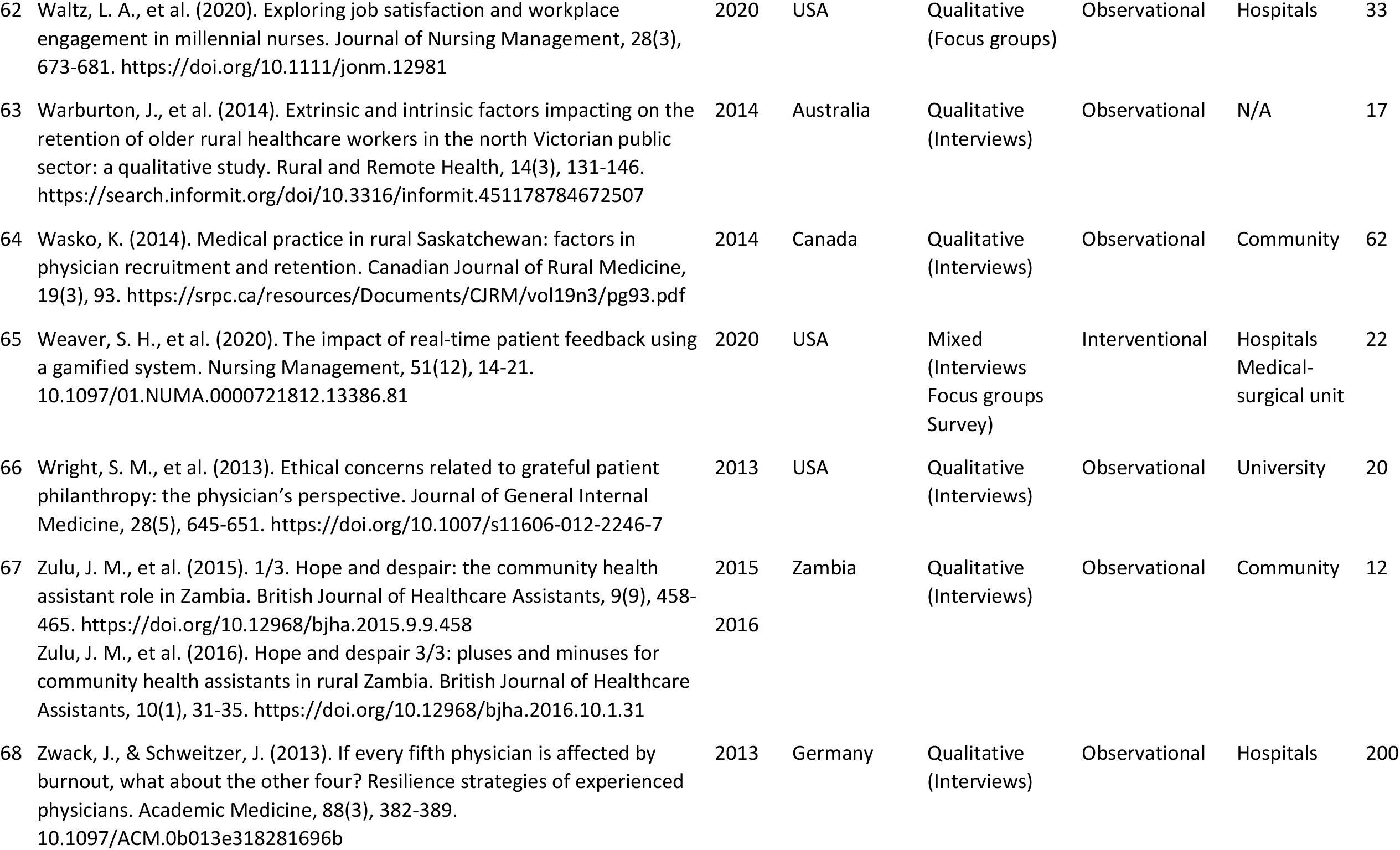
Summary data abstraction table.

| ID | Reference | Year | Country | Study type | Design | Setting | Sample size |
| --- | --- | --- | --- | --- | --- | --- | --- |
| 1 | Akintola, O. (2010). Perceptions of rewards among volunteer caregivers of people living with AIDS working in faith-based organizations in South Africa: a qualitative study. *Journal of the International AIDS Society*, 13(1), 1-10. https://doi.org/10.1186/1758-2652-13-22 | 2010 | South Africa | Qualitative (Interviews) | Observational | Community | 55 |
| 2 | Akintola, O., & Chikoko, G. (2016). Factors influencing motivation and job satisfaction among supervisors of community health workers in marginalized communities in South Africa. *Human Resources for Health*, 14(1), 1-15. https://doi.org/10.1186/s12960-016-0151-6 | 2016 | South Africa | Qualitative (Interviews) | Observational | Community | 26 |
| 3 | Alam, K., & Oliveras, E. (2014). Retention of female volunteer community health workers in Dhaka urban slums: a prospective cohort study. *Human Resources for Health*, 12(1), 1-11. https://doi.org/10.1186/1478-4491-12-29 | 2014 | Bangladesh | Mixed (Interviews Survey) | Observational | Community | 542 |
| 4 | Alam, K., et al. (2012). Performance of female volunteer community health workers in Dhaka urban slums. *Social Science & Medicine*, 75(3), 511-515. https://doi.org/10.1016/j.socscimed.2012.03.039 | 2012 | Bangladesh | Mixed (Questionnaire Focus groups Interviews) | Observational | Community | 542 |
| 5 | Alibhai, A. A. (2013). The effectiveness of a volunteer community health worker program to support an antiretroviral treatment program for AIDS patients in western Uganda. Available from ProQuest Dissertations & Theses A&I. | 2013 | Uganda | Mixed (Questionnaire Interviews Focus groups) | Observational | Community | 169 |
| 6 | Aparicio, M., et al. (2019). Gratitude from patients and relatives in palliative care—characteristics and impact: a national survey. *BMJ Supportive & Palliative Care*. 10.1136/bmjspcare-2019-001858 | 2019 | Spain | Quantitative (Survey) | Observational | Palliative care units<br>Community | 186 |
| 7 | Ashley, C., et al. (2021). The psychological well-being of primary healthcare nurses during COVID-19: a qualitative study. *Journal of Advanced Nursing*, 77(9), 3820-3828. https://doi.org/10.1111/jan.14937 | 2021 | Australia | Qualitative (Interviews) | Observational | GPs<br>Community | 25 |
| 8 | Bakker, D., et al. (2010). Canadian cancer nurses' views on recruitment and retention. *Journal of Nursing Management*, 18(2), 205-214. https://doi.org/10.1111/j.1365-2834.2009.01029.x | 2010 | Canada | Qualitative (Focus groups) | Observational | Oncology<br>Ambulatory care<br>Hospitals<br>Community | 91 |
| 9 | Barnes, A. L. (2015). Relationship between job satisfaction among frontline staff and patient satisfaction: Evidence from community health centers in South Carolina (Doctoral dissertation, University of South Carolina). https://www.proquest.com/docview/1765406972?pq-origsite=gscholar&fromopenview=true | 2015 | USA | Quantitative (Survey) | Observational | Community | 303 |
| 10 | Beate, A., & Jacobsen, F. F. (2020). The art of caring in selected Norwegian nursing homes: a qualitative approach. *International Journal of Caring Sciences*, 13(2), 820. https://hdl.handle.net/11250/2738332 | 2020 | Norway | Qualitative (Interviews) | Observational | Nursing homes | 11 |
| 11 | Bhatnagar, A. (2014). Determinants of motivation and job satisfaction among primary health workers: case studies from Nigeria and India (Doctoral dissertation, Johns Hopkins University). http://jhir.library.jhu.edu/handle/1774.2/37851 | 2014 | Nigeria<br>India | Mixed (Interviews<br>Survey) | Observational | Primary health care | 29 |
| 12 | Bhatnagar, A., et al. (2017). Primary health care workers' views of motivating factors at individual, community and organizational levels: a qualitative study from Nasarawa and Ondo states, Nigeria. *The International Journal of Health Planning and Management*, 32(2), 217-233. https://doi.org/10.1002/hpm.2342 | 2017 | Nigeria | Qualitative (Interviews) | Observational | Community | 29 |
| 13 | Blank, F. S., et al. (2014). A comparison of patient and nurse expectations regarding nursing care in the emergency department. *Journal of Emergency Nursing*, 40(4), 317-322. https://doi.org/10.1016/j.jen.2013.02.010 | 2014 | N/A | Mixed (Survey) | Observational | Emergency department | 100 |
| 14 | Cameron, P. J., et al. (2010). Physician retention in rural Alberta: key community factors. *Canadian Journal of Public Health*, 101(1), 79-82. https://doi.org/10.1007/BF03405568 | 2010 | Canada | Qualitative (Interviews<br>Document review<br>Observations) | Observational | Community | 15 |
| 15 | Chou, W. C., et al. (2006). Perceptions of physicians on the barriers and facilitators to integrating fall risk evaluation and management into practice. *Journal of General Internal Medicine*, 21(2), 117-122. https://doi.org/10.1007/s11606-006-0244-3 | 2006 | USA | Qualitative (Interviews) | Observational | Primary care offices | 18 |
| 16 | Christiansen, B. (2008). Good work—how is it recognised by the nurse? *Journal of Clinical Nursing*, 17(12), 1645-1651. https://doi.org/10.1111/j.1365-2702.2007.02139.x | 2008 | Norway | Qualitative (Interviews) | Observational | Hospitals<br>Clinic | 10 |
| 17 | Ciocănel, A., et al. (2018). Helping, mediating, and gaining recognition: the everyday identity work of Romanian health social workers. *Social Work in Health Care*, 57(3), 206-219. https://doi.org/10.1080/00981389.2018.1426674 | 2018 | Romania | Qualitative (Interviews) | Observational | Hospitals<br>Emergency department<br>Maternity unit<br>School-based<br>Community<br>Hospice | 21 |
| 18 | Cleary, M., et al. Mental health nurses' perceptions of good work in an acute setting. *International Journal of Mental Health Nursing*, 21(5), 471-479. https://doi.org/10.1111/j.1447-0349.2011.00810.x | 2012 | Australia | Qualitative (Interviews) | Observational | Mental health centres | 40 |
| 19 | Converso, D., et al. (2015). Do positive relations with patients play a protective role for healthcare employees? Effects of patients' gratitude and support on nurses' burnout. *Frontiers in Psychology*, 6, 470. [10.3389/fpsyg.2015.00470](https://doi.org/10.3389/fpsyg.2015.00470) | 2015 | Italy | Quantitative (Questionnaire) | Observational | Hospitals<br>Emergency department<br>Oncology | 204 |
| 20 | Cortese, C. G. (2007). Job satisfaction of Italian nurses: an exploratory study. *Journal of Nursing Management*, 15(3), 303-312. https://doi.org/10.1111/j.1365-2834.2007.00694.x | 2007 | Italy | Qualitative (Interviews) | Observational | Hospitals | 64 |
| 21 | Dageid, W., et al. (2016). Sustaining motivation among community health workers in aids care in Kwazulu-natal, South Africa: challenges and prospects. *Journal of Community Psychology*, 44(5), 569-585. https://doi.org/10.1002/jcop.21787 | 2016 | South Africa | Qualitative (Interviews) | Observational | Community | 12 |
| 22 | Danet, A. D., et al. (2020). Emotional paths of professional experiences in transplant coordinators. *Nefrología (English Edition)*, 40(1), 75-90. https://doi.org/10.1016/j.nefro.2019.05.005 | 2020 | Spain | Qualitative (Questionnaire Interviews) | Observational | Hospitals<br>Transplant coordination | 22 |
| 23 | Datiko, D. G., et al. (2015). Exploring providers' perspectives of a community based TB approach in Southern Ethiopia: implication for community based approaches. *BMC Health Services Research*, 15(1), 1-9. https://doi.org/10.1186/s12913-015-1149-9 | 2015 | Ethiopia | Qualitative (Interviews) | Observational | Community | 37 |
| 24 | de Oliveira, A. R., et al. (2019). Satisfaction and limitation of primary health care nurses' work in rural areas. *Rural and Remote Health*, 19(2), 55-64. https://search.informit.org/doi/10.3316/informit.143753391883465 | 2019 | Brazil | Qualitative (Interviews) | Observational | Family health units | 11 |
| 25 | Fereday, J., & Muir-Cochrane, E. (2006). The role of performance feedback in the self-assessment of competence: a research study with nursing clinicians. *Collegian*, 13(1), 10-15. [https://doi.org/10.1016/S1322-7696(08)60511-9](https://doi.org/10.1016/S1322-7696%2808%2960511-9) | 2006 | Australia | Qualitative (Focus groups) | Observational | Hospitals<br>Midwifery<br>General surgical<br>General medical | 26 |
| 26 | Fontanini, R., et al. (2021). Italian nurses' experiences during the COVID-19 pandemic: a qualitative analysis of internet posts. *International Nursing Review*, 68(2), 238-247. https://doi.org/10.1111/inr.12669 | 2021 | Italy | Qualitative (Descriptive study) | Observational | Hospitals<br>Community | 380 |
| 27 | Fort, A. L., & Voltero, L. (2004). Factors affecting the performance of maternal health care providers in Armenia. *Human Resources for Health*, 2(1), 1-11. https://doi.org/10.1186/1478-4491-2-8 | 2004 | Armenia | Quantitative (Interviews Survey Observations) | Observational | Reproductive health services | 285 |
| 28 | Johansson, M., et al. (2019). Nursing staff's experiences of intensive care unit diaries: a qualitative study. *Nursing in Critical Care*, 24(6), 407-413. https://doi.org/10.1111/nicc.12416 | 2019 | Sweden | Qualitative (Focus groups) | Observational | University Hospitals<br>ICU | 27 |
| 29 | Judd, M. J., et al. (2017). Workplace stress, burnout and coping: a qualitative study of the experiences of Australian disability support workers. *Health & Social Care in the Community*, 25(3), 1109-1117. https://doi.org/10.1111/hsc.12409 | 2017 | Australia | Qualitative (Interviews) | Observational | Disability Services | 12 |
| 30 | Kelly, D., et al. (2020). The experiences of cancer nurses working in four European countries: a qualitative study. *European Journal of Oncology Nursing*, 49, 101844. https://doi.org/10.1016/j.ejon.2020.101844 | 2020 | Estonia<br>Germany<br>Netherlands<br>UK | Qualitative (Interviews Focus groups) | Observational | Oncology | 97 |
| 31 | Khowaja, K., et al. (2005). Registered nurses perception of work satisfaction at a Tertiary Care University Hospital. *Journal of Nursing Management*, 13(1), 32-39. https://doi.org/10.1111/j.1365-2834.2004.00507.x | 2005 | Pakistan | Qualitative (Interviews Focus groups) | Observational | Hospitals<br>Critical care<br>Medical-surgery<br>Ambulatory<br>Maternity<br>Emergency department | 45 |
| 32 | Kim, Y. M., et al. (2008). Factors that enable nurse-patient communication in a family planning context: a positive deviance study. *International Journal of Nursing Studies*, 45(10), 1411-1421. https://doi.org/10.1016/j.ijnurstu.2008.01.002 | 2008 | Indonesia | Qualitative (Interviews Focus groups) | Observational | Clinic | 34 |
| 33 | MacLeod, M. L., et al. (2021). The meaning of nursing practice for nurses who are retired yet continue to work in a rural or remote community. *BMC Nursing*, 20(1), 1-13. https://doi.org/10.1186/s12912-021-00721-0 | 2021 | Canada | Qualitative (Survey) | Observational | N/A | 101 |
| 34 | Maharani, C., et al. (2022). The National Health Insurance System of Indonesia and primary care physicians' job satisfaction: a prospective qualitative study. *Family Practice*, 39(1), 112–124. https://doi.org/10.1093/fampra/cmab067 | 2022 | Indonesia | Qualitative (Interviews) | Observational | Primary health care | 34 |
| 35 | Martínez-Taboas, A., et al. (2014). Gifts in psychotherapy: attitudes and experiences of Puerto Rican psychotherapists. *Revista Puertorriqueña de Psicología*, 25(2), 328-339. https://www.redalyc.org/articulo.oa?id=233245622011 | 2014 | Puerto Rico | Quantitative (Questionnaire) | Observational | Private practice Hospitals University | 75 |
| 36 | Minooee, S., et al. (2021). Catastrophic thinking: is it the legacy of traumatic births? Midwives' experiences of shoulder dystocia complicated births. *Women and Birth*, 34(1), e38-e46. https://doi.org/10.1016/j.wombi.2020.08.008 | 2021 | Australia | Qualitative (Interviews) | Observational | Hospitals | 25 |
| 37 | Muntz, J., & Dormann, C. (2020). Moderating effects of appreciation on relationships between illegitimate tasks and intrinsic motivation: a two-wave shortitudinal study. *European Journal of Work and Organizational Psychology*, 29(3), 391-404. https://doi.org/10.1080/1359432X.2019.1706489 | 2020 | Germany | Quantitative (Panel study) | Observational | Hospitals | 241 |
| 38 | Nwala, E. (2015). The impact of nonmonetary job benefits on job retention in rural healthcare (Doctoral dissertation, Capella University). https://www.proquest.com/docview/1735405605?pq-origsite=gscholar&fromopenview=true | 2015 | USA | Qualitative (Interviews Observations) | Observational | Clinic | 13 |
| 39 | Oluwole, A., et al. (2019). Optimising the performance of frontline implementers engaged in the NTD programme in Nigeria: lessons for strengthening community health systems for universal health coverage. *Human Resources for Health*, 17(1), 1-16. https://doi.org/10.1186/s12960-019-0419-8 | 2019 | Nigeria | Qualitative (Workshops) | Observational | Community | N/A |
| 40 | Ortiz, J. A. (2014). New graduate nurses' experiences of what accounts for their lack of professional confidence during their first year of practice (Doctoral dissertation, Capella University). https://www.proquest.com/docview/1650654883?pq-origsite=gscholar&fromopenview=true | 2014 | USA | Qualitative (Interviews) | Observational | Hospitals | 12 |
| 41 | Pal, L. M., et al. (2014). Utilising feedback from patients and their families as a learning strategy in a foundation degree in palliative and supportive care: a qualitative study. *Nurse Education Today*, 34(3), 319-324. https://doi.org/10.1016/j.nedt.2013.06.012 | 2014 | UK | Qualitative (Focus groups Questionnaire) | Observational | Nursing homes Hospitals Hospices Oncology wards Community | 12 |
| 42 | Pariseault, C. A., et al. (2022). Nurses' experiences of caring for patients and families during the Covid-19 pandemic: communication challenges. *American Journal of Nursing*, 122, 22-30. https://doi.org/10.1097/01.NAJ.0000805644.85184.d2 | 2022 | USA | Qualitative (Descriptive study) | Observational | Hospitals | 17 |
| 43 | Peteet, J. R., et al. (1992). Relationships with patients in oncology: can a clinician be a friend? *Psychiatry*, 55(3), 223-229. https://doi.org/10.1080/00332747.1992.11024596 | 1992 | USA | Mixed (Interviews) | Observational | Oncology | 192 |
| 44 | Pooley, H. M., et al. (2015). The experience of the long-term doctor-patient relationship in consultant nephrologists. *Journal of Renal Care*, 41(2), 88-95. https://doi.org/10.1111/jorc.12092 | 2015 | UK | Qualitative (Interviews) | Observational | Renal department | 7 |
| 45 | Prytherch, H., et al. (2012). Maternal and newborn healthcare providers in rural Tanzania: in-depth interviews exploring influences on motivation, performance and job satisfaction. *Rural and Remote Health*, 12(3), 1-15. https://search.informit.org/doi/10.3316/informit.625974688045681 | 2012 | Tanzania | Qualitative (Interviews) | Observational | Health centres | 35 |
| 46 | Prytherch, H., et al. (2013). Motivation and incentives of rural maternal and neonatal health care providers: a comparison of qualitative findings from Burkina Faso, Ghana and Tanzania. *BMC Health Services Research*, 13(1), 1-15. https://doi.org/10.1186/1472-6963-13-149 | 2013 | Burkina Faso<br>Ghana<br>Tanzania | Qualitative (Interviews) | Observational | Health centres | 35 |
| 47 | Raingruber, B., & Wolf, T. (2015). Nurse perspectives regarding the meaningfulness of oncology nursing practice. *Clinical Journal of Oncology Nursing*, 19(3), 292-296. [10.1188/15.CJON.292-296](https://doi.org/10.1188/15.CJON.292-296) | 2015 | USA | Qualitative (Interviews) | Observational | Oncology wards<br>Medical-surgical unit | 8 |
| 48 | Reis, M. J. D., et al. (2010). Experiences of nurses in health care for female victims of sexual violence. *Revista de Saude Publica*, 44, 325-331. https://doi.org/10.1590/S0034-89102010000200013 | 2010 | Brazil | Qualitative (Interviews) | Observational | Sexual violence service | 6 |
| 49 | Riskin, A., et al. (2019). Expressions of gratitude and medical team performance. *Pediatrics*, 143(4). https://doi.org/10.1542/peds.2018-2043 | 2019 | Israel | Quantitative (Randomised study) | Interventional | Hospitals NICU | 172 |
| 50 | Robinson, D. (2019). Exploring experiences of burnout, engagement, and social support networks: a qualitative study of hospital medicine physicians (Doctoral dissertation, Colorado State University). https://www.proquest.com/docview/2244361153?pq-origsite=gscholar&fromopenview=true | 2019 | USA | Mixed (Interviews Survey) | Observational | Hospitals | 15 |
| 51 | Roca, J., et al. (2021). Experiences, emotional responses, and coping skills of nursing students as auxiliary health workers during the peak Covid-19 pandemic: a qualitative study. *International Journal of Mental Health Nursing*, 30(5), 1080-1092. https://doi.org/10.1111/inm.12858 | 2021 | Spain | Qualitative (Interviews) | Observational | Nursing homes<br>Hospitals<br>COVID-19 specialized unit | 22 |
| 52 | Ronnie, L. (2019). Intensive care nurses in South Africa: expectations and experiences in a public sector hospital. *Journal of Nursing Management*, 27(7), 1431-1437. https://doi.org/10.1111/jonm.12826 | 2019 | South Africa | Qualitative (Interviews) | Observational | Hospitals ICU | 44 |
| 53 | Sakai, M., et al. (2013). Home visiting nurses' attitudes toward caring for dying patients, and related workplace factors. *International Journal of Palliative Nursing*, 19(4), 195-204. https://doi.org/10.12968/ijpn.2013.19.4.195 | 2013 | Japan | Quantitative (Questionnaire) | Observational | Community | 206 |
| 54 | Seitovirta, J., et al. (2015). Registered nurses' experiences of rewarding in a Finnish university hospital—an interview study. *Journal of Nursing Management*, 23(7), 868-878. https://doi.org/10.1111/jonm.12228 | 2015 | Finland | Qualitative (Interviews) | Observational | Hospitals | 10 |
| 55 | Seitovirta, J., et al. (2017). Attention to nurses' rewarding—an interview study of registered nurses working in primary and private healthcare in Finland. *Journal of Clinical Nursing*, 26(7-8), 1042-1052. https://doi.org/10.1111/jocn.13459 | 2017 | Finland | Qualitative (Interviews) | Observational | Healthcare organisations | 20 |
| 56 | Smallwood, N., et al. (2021). Moral distress and perceived community views are associated with mental health symptoms in frontline health workers during the COVID-19 pandemic. *International Journal of Environmental Research and Public Health*, 18(16), 8723. https://doi.org/10.3390/ijerph18168723 | 2021 | Australia | Quantitative (Survey) | Observational | Hospitals | 7846 |
| 57 | Tang, P. M., et al. (2021). How and when service beneficiaries' gratitude enriches employees' daily lives. *Journal of Applied Psychology*, 107(6), 987-1008. https://doi.org/10.1037/apl0000975 | 2021 | China<br>Singapore | Quantitative (Experience Sampling Method) | Observational | Hospitals | 275 |
| 58 | Vachon, M., & Guité-Verret, A. (2020). From powerlessness to recognition the meaning of palliative care clinicians' experience of suffering. *International Journal of Qualitative Studies on Health and Well-being*, 15(1), 1852362. https://doi.org/10.1080/17482631.2020.1852362 | 2020 | Canada | Qualitative (Interviews) | Observational | Medical centre | 21 |
| 59 | Vail, L., et al. (2011). Healthcare assistants in general practice: a qualitative study of their experiences. *Primary Health Care Research & Development*, 12(1), 29-41. https://doi.org/10.1017/S1463423610000204 | 2011 | UK | Qualitative (Interviews) | Observational | GP | 14 |
| 60 | Vandecasteele, T., et al. (2015). Nurses' perceptions of transgressive behaviour in care relationships: a qualitative study. *Journal of Advanced Nursing*, 71(12), 2786-2798. https://doi.org/10.1111/jan.12749 | 2015 | Belgium | Qualitative (Interviews) | Observational | Hospitals | 18 |
| 61 | Wahlberg, A. C., & Bjorkman, A. (2018). Expert in nursing care but sometimes disrespected—telenurses' reflections on their work environment and nursing care. *Journal of Clinical Nursing*, 27(21-22), 4203-4211. https://doi.org/10.1111/jocn.14622 | 2018 | Sweden | Qualitative (Interviews) | Observational | Telephone service | 24 |
| 62 | Waltz, L. A., et al. (2020). Exploring job satisfaction and workplace engagement in millennial nurses. *Journal of Nursing Management*, 28(3), 673-681. https://doi.org/10.1111/jonm.12981 | 2020 | USA | Qualitative (Focus groups) | Observational | Hospitals | 33 |
| 63 | Warburton, J., et al. (2014). Extrinsic and intrinsic factors impacting on the retention of older rural healthcare workers in the north Victorian public sector: a qualitative study. *Rural and Remote Health*, 14(3), 131-146. https://search.informit.org/doi/10.3316/informit.451178784672507 | 2014 | Australia | Qualitative (Interviews) | Observational | N/A | 17 |
| 64 | Wasko, K. (2014). Medical practice in rural Saskatchewan: factors in physician recruitment and retention. *Canadian Journal of Rural Medicine*, 19(3), 93. https://srpc.ca/resources/Documents/CJRM/vol19n3/pg93.pdf | 2014 | Canada | Qualitative (Interviews) | Observational | Community | 62 |
| 65 | Weaver, S. H., et al. (2020). The impact of real-time patient feedback using a gamified system. *Nursing Management*, 51(12), 14-21. 10.1097/01.NUMA.0000721812.13386.81 | 2020 | USA | Mixed (Interviews Focus groups Survey) | Interventional | Hospitals Medical-surgical unit | 22 |
| 66 | Wright, S. M., et al. (2013). Ethical concerns related to grateful patient philanthropy: the physician's perspective. *Journal of General Internal Medicine*, 28(5), 645-651. https://doi.org/10.1007/s11606-012-2246-7 | 2013 | USA | Qualitative (Interviews) | Observational | University | 20 |
| 67 | Zulu, J. M., et al. (2015). 1/3. Hope and despair: the community health assistant role in Zambia. *British Journal of Healthcare Assistants*, 9(9), 458-465. https://doi.org/10.12968/bjha.2015.9.9.458<br>Zulu, J. M., et al. (2016). Hope and despair 3/3: pluses and minuses for community health assistants in rural Zambia. *British Journal of Healthcare Assistants*, 10(1), 31-35. https://doi.org/10.12968/bjha.2016.10.1.31 | 2015<br>2016 | Zambia | Qualitative (Interviews) | Observational | Community | 12 |
| 68 | Zwack, J., & Schweitzer, J. (2013). If every fifth physician is affected by burnout, what about the other four? Resilience strategies of experienced physicians. *Academic Medicine*, 88(3), 382-389. 10.1097/ACM.0b013e318281696b | 2013 | Germany | Qualitative (Interviews) | Observational | Hospitals | 200 |

Research was located in 32 countries across six continents (Table 2). Two studies were located in multiple countries (50, 51). One study did not state the study location (52).

**Table 2.** Research location of included studies in order of quantity. Multiple papers from the same study counted as having a signal location unless reporting results fro different locations.

| Continent | Quantity | Country | Quantity | Study ID(s) |
| --- | --- | --- | --- | --- |
| Europe | 23 | UK | 4 | 30, 41, 44, 59 |
|  |  | Germany | 3 | 30, 37, 68 |
|  |  | Italy | 3 | 19, 20, 26 |
|  |  | Spain | 3 | 6, 22, 51 |
|  |  | Finland | 2 | 54, 55 |
|  |  | Norway | 2 | 10, 16 |
|  |  | Sweden | 2 | 28, 61 |
|  |  | Belgium | 1 | 60 |
|  |  | Estonia | 1 | 30 |
|  |  | Netherlands | 1 | 30 |
|  |  | Romania | 1 | 17 |
| North America | 16 | USA | 11 | 9, 15, 38, 40, 42, 43, 47, 50, 62, 65, 66 |
|  |  | Canada | 5 | 8, 14, 33, 58, 64 |
| Africa | 12 | South Africa | 4 | 1, 2, 21, 52 |
|  |  | Nigeria | 2 | 11, 39 |
|  |  | Burkina Faso | 1 | 46 |
|  |  | Ethiopia | 1 | 23 |
|  |  | Ghana | 1 | 46 |
|  |  | Tanzania | 1 | 46 |
|  |  | Uganda | 1 | 5 |
|  |  | Zambia | 1 | 67 |
| Asia | 9 | Indonesia | 2 | 32, 34 |
|  |  | Armenia | 1 | 27 |
|  |  | Bangladesh | 1 | 4 |
|  |  | China | 1 | 57 |
|  |  | Israel | 1 | 49 |
|  |  | Japan | 1 | 53 |
|  |  | Pakistan | 1 | 31 |
|  |  | Singapore | 1 | 57 |
| Australasia | 7 | Australia | 7 | 7, 18, 25, 29, 36, 56, 63 |
| South America | 3 | Brazil | 2 | 24, 48 |
|  |  | Puerto Rico | 1 | 35 |

The median year of publication was 2015 (Table 3).

**Table 3.** Year of publication for included papers in chronological order with corresponding study IDs. Multiple papers from the same study were included separately due to differing publication dates

| Year | Quantity | Study ID(s) |
| --- | --- | --- |
| 1992 | 1 | 43 |
| 2004 | 1 | 27 |
| 2005 | 1 | 31 |
| 2006 | 2 | 15, 25 |
| 2007 | 1 | 20 |
| 2008 | 2 | 16, 32 |
| 2010 | 4 | 1, 8, 14, 48 |
| 2011 | 1 | 59 |
| 2012 | 3 | 4, 18, 45 |
| 2013 | 5 | 8, 11, 32, 34, 45 |
| 2014 | 8 | 3, 11, 13, 35, 40, 41, 63, 64 |
| 2015 | 8 | 9, 19, 23, 38, 44, 47, 54, 60 |
| 2016 | 3 | 2, 21, 67 |
| 2017 | 3 | 12, 29, 5 |
| 2018 | 2 | 17, 61 |
| 2019 | 7 | 6, 24, 28, 39, 49, 50, 52 |
| 2020 | 7 | 10, 22, 30, 37, 58, 62, 65 |
| 2021 | 7 | 7, 26, 33, 36, 51, 56, 57 |
| 2022 | 2 | 34, 42 |

Most studies were qualitative, and all but two studies were observational, in that they presented evidence relating to existing uses of positive feedback (Table 4).

**Table 4.** Methods of included papers, in order of quantity. Multiple papers from the same study were counted as having a single study methods. Three companion papers were not counted in the ‘total quantity’ column. Many papers used multiple methods, each counted separately in the ‘quantity’ column.

| Type of study | Total quantity | Method | Quantity |
| --- | --- | --- | --- |
| Qualitative | 49 | Interviews | 40 |
|  |  | Focus groups | 8 |
|  |  | Questionnaire/survey | 4 |
|  |  | Observations | 2 |
|  |  | Descriptive study | 2 |
|  |  | Workshops | 1 |
| Quantitative | 10 | Questionnaire/survey | 6 |
|  |  | Experience Sampling Method | 1 |
|  |  | Observations | 1 |
|  |  | Panel study | 1 |
|  |  | Randomised study | 1 |
| Mixed | 6 | Interviews | 5 |
|  |  | Questionnaire/survey | 5 |
|  |  | Focus groups | 3 |

The two intervention studies were as follows:

#### Riskin et al, 2019 [ID 49]

This study used pre-recorded video to simulate the impact on Neonatal Intensive Care Unit (NICU) team performance of gratitude expressed by two different sources. NICU teams (n = 43) were randomly assigned to 1 of 4 conditions: (1) maternal gratitude (2) physician-expressed gratitude (3) combined maternal and physician gratitude, or (4) control (same agents communicated neutral statements). Subsequent team performance in a training workshop was evaluated by a blinded panel, on a five-point Likert scale. Maternal gratitude produced a significant positive affect on team performance (3.9 ± 0.9 vs 3.6 ± 1.0; P = .04). Most of this effect was explained by the positive impact of gratitude on team information sharing (4.3 ± 0.8 vs 4.0 ± 0.8; P = .03). As a result, accuracy of diagnostic work was improved (95% CI = 0.018 to 0.395; *P* = .03)

#### Weaver, 2020 [ID 65]

This study evaluated the impact of using a gamified feedback system on a medical-surgical unit in the US. The feedback system allowed service users to use a tablet to input free-text comments, which were later sent as text alerts to nurses and technicians. Its impact was evaluated using interviews, focus groups, and surveys. Healthcare staff described that receiving recognition and appreciation through the feedback system made them feel good, boosted confidence, morale and motivation, and helped them to feel comfortable in their job. Staff were initially enthusiastic about using the feedback system, which was seen to support the effect of positive feedback. Similarly, when staff became less enthusiastic and motivated to use the system over time, this hindered the effects of positive feedback. Night shift staff reported less opportunity to receive feedback from service users. The system was hindered by the lengthy process of accumulating points and rewards, making feedback from service users less timely, consistent, or meaningful.

### Characteristics of positive feedback in included studies

Positive feedback was described in included studies as having a variety of forms, most commonly described in their original papers as appreciation and gratitude (Table 5). The form of feedback was categorised as material or ambiguous. Material feedback referred to physical items given by service users, families, or the community. In a substantial number of included papers, the precise form of feedback was not explicitly stated, and hence was ambiguous. For example, gratitude might incorporate a variety of actions, but this was often not stated in published work.

**Table 5.** Positive feedback in included studies in order of quantity. Multiple papers from the same study were counted as having a single type of feedback

| Feedback category | Type of positive feedback | Quantity | Study ID(s) |
| --- | --- | --- | --- |
| Ambiguous | Appreciation | 28 | 1, 2, 7, 8, 9, 11, 14, 19, 21, 24, 26, 29, 32, 33, 34, 36, 37, 43, 45, 47, 49, 51, 53, 54, 56, 61, 64, 68 |
|  | Gratitude | 22 | 6, 10, 13, 14, 20, 21, 23, 24, 32, 33, 35, 42, 43, 46, 48, 52, 53, 57, 59, 60, 61, 62 |
|  | Thanks | 16 | 5, 6, 14, 18, 30, 33, 35, 36, 40, 46, 47, 50, 54, 55, 65, 66 |
|  | Positive feedback | 13 | 4, 15, 16, 17, 25, 28, 31, 38, 39, 40, 41, 44, 62 |
|  | Recognition | 10 | 1, 11, 24, 27, 32, 43, 50, 55, 60, 68 |
|  | Praise | 3 | 28, 41, 50 |
|  | Being valued | 1 | 63 |
|  | Patient satisfaction | 1 | 67 |
| Material | Gifts | 7 | 6, 14, 22, 25, 52, 58, 66 |
|  | Cards | 5 | 14, 16, 18, 25, 65 |
|  | Flowers | 2 | 52, 65 |
|  | Food | 2 | 6, 62 |
|  | Hugs | 1 | 16 |
|  | Letters | 1 | 6 |

Included studies identified that positive feedback was delivered by service users (n = 53), the community (n = 18), and families (n = 16), with some studies identifying multiple sources of feedback.

Recipients of positive feedback were described using a broad variety of labels, most commonly identified as clinical staff providing direct care and treatment to service users (n = 68) (Table 6). In some studies, non-clinical staff received feedback (n = 3).

**Table 6.** Feedback recipients of positive feedback in included studies in order of quantity. Multiple papers from the same study were counted as a single feedback recipient

| Recipient category | Feedback recipient | Quantity | Study ID(s) |
| --- | --- | --- | --- |
| Clinical staff | Nurses | 29 | 7, 8, 13, 16, 18, 19, 20, 24, 25, 26, 27, 28, 31, 32, 33, 37, 40, 42, 47, 48, 52, 53, 54, 55, 57, 60, 61, 62, 65 |
|  | Community health workers | 7 | 4, 5, 7, 21, 23, 39, 67 |
|  | Physicians | 6 | 14, 15, 43, 64, 66, 68 |
|  | Healthcare professionals | 3 | 6, 22, 34 |
|  | Clinical staff | 2 | 30, 58 |
|  | Frontline health workers | 2 | 9, 56 |
|  | Health social workers | 2 | 17, 43 |
|  | Healthcare personnel | 2 | 10, 38 |
|  | Healthcare students | 2 | 41, 51 |
|  | Midwives | 2 | 27, 36 |
|  | Adult treatment team members | 1 | 43 |
|  | Doctors | 1 | 57 |
|  | Healthcare assistant | 1 | 59 |
|  | Healthcare providers | 1 | 45 |
|  | Healthcare workers | 1 | 63 |
|  | Hospitalists | 1 | 50 |
|  | Neonatal Intensive Care Unit team | 1 | 49 |
|  | Nephrologists | 1 | 44 |
|  | Primary health worker | 1 | 11 |
|  | Psychologists | 1 | 35 |
|  | Volunteer community caregivers | 1 | 1 |
| Non-clinical staff | Supervisors | 1 | 2 |
|  | Technicians | 1 | 65 |
|  | Disability support worker | 1 | 29 |

Healthcare staff worked in a range of settings, categorised as clinical (primarily provides a health-related medical function) and non-clinical (primary purpose is not to provide a direct health-related medical function). Most studies considered clinical settings (n = 74) (Table 7). Two included papers did not explicitly state the setting (53, 54).

**Table 7.** Feedback settings of positive feedback delivery in included studies in order of quantity. Multiple papers from the same study were counted separately only if the setting differed between papers

| Setting category | Feedback setting | Quantity | Study ID(s) |
| --- | --- | --- | --- |
| Clinical setting | Hospitals | 27 | 8, 16, 17, 19, 20, 22, 25, 26, 28, 31, 35, 36, 37, 40, 41, 42, 49, 50, 51, 52, 54, 56, 57, 60, 62, 65, 68 |
|  | Oncology | 6 | 8, 19, 30, 41, 43, 47 |
|  | Emergency department | 4 | 13, 17, 19, 31 |
|  | Clinics | 3 | 16, 32, 38 |
|  | General Practice (GP) | 3 | 7, 25, 59 |
|  | Health centres | 3 | 18, 45, 58 |
|  | Intensive care | 3 | 28, 49, 52 |
|  | Maternal care | 3 | 17, 25, 31 |
|  | Medical surgery | 3 | 31, 47, 65 |
|  | Nursing homes | 3 | 10, 41, 51 |
|  | Primary care | 3 | 11, 15, 34 |
|  | Ambulatory care | 2 | 8, 31 |
|  | Hospices | 2 | 17, 41 |
|  | Covid-19 unit | 1 | 51 |
|  | Critical care | 1 | 31 |
|  | Family health units | 1 | 24 |
|  | Palliative care | 1 | 6 |
|  | Private practice | 1 | 35 |
|  | Renal department | 1 | 44 |
|  | Reproductive health services | 1 | 27 |
|  | Sexual violence services | 1 | 48 |
|  | Transplant coordination | 1 | 22 |
| Non-clinical setting | Community (including home-based care and faith-based organisations) | 19 | 1, 2, 4, 5, 6, 7, 8, 9, 12, 14, 17, 21, 23, 26, 39, 41, 53, 64, 67 |
|  | University | 3 | 28, 35, 66 |
|  | Disability services | 1 | 29 |
|  | School-based | 1 | 17 |
|  | Telephone services | 1 | 61 |

### Objective 1: Measures of change

There was a considerable variation in the outcome domains and measures used in studies (n = 11) (Table 8). The remaining 57 studies did not include a standardised outcome measure. A measure was concluded to be standardised if a citable reference was available.

**Table 8.** Outcome domains and outcome measures used in included studies.

| Outcome domain | Standardised outcome measure | Quantity | Study ID(s) |
| --- | --- | --- | --- |
| Attitudes towards caring for dying patients | Frommelt Attitudes Toward Care of the Dying scale, form B, Japanese version (FATCOD B-J) | 1 | 53 |
| Attitudes towards death | The Death Attitude Inventory (DAI) | 1 | 53 |
| Baseline affective states | Short Positive and Negative Affect Schedule (PANAS) | 1 | 57 |
| Beliefs, attitudes, experiences of gifts | Scale of Attitudes and Behaviors toward Gifts in Psychotherapy (SABGP) | 1 | 35 |
| Burnout | Maslach Burnout Inventory | 2 | 50, 68 |
| Burnout | Maslach Burnout Inventory for Human Service Sector | 1 | 19 |
| Completion of clinical/non-clinical tasks | MEASURE Evaluation's Quick Investigation of Quality (QIQ) tool | 1 | 27 |
| Engagement at work | Gallup Worker Engagement Survey | 2 | 50, 65 |
| Experiences, understandings, meanings | Nursing Practice in Rural and Remote Canada II (RRNII) | 1 | 33 |
| Illegitimate tasks | Bern Illegitimate Tasks Scale | 1 | 37 |
| Job satisfaction | Job Enjoyment Scale | 1 | 65 |
| Patient behaviour as a psychological resource | Customer-initiated support scale | 1 | 19 |
| Patient satisfaction | Hospital Consumer Assessment of Health Plans Survey (HCAHPS) | 1 | 65 |
| Perception of service user gratitude | PGRate scale | 1 | 19 |
| Psychological demands | Job Content Questionnaire (JCQ) subscales | 1 | 19 |
| Resilience | Abbreviated 2 item Connor-Davidson Resilience Scale (CD-RISC 2) | 1 | 56 |
| Resilience | Abbreviated Impact of Event Scale (IES-6) | 1 | 56 |
| Resilience | Abbreviated Maslach Burnout Inventory (AMBI) | 1 | 56 |
| Resilience | Patient Health Questionnaire (PHQ-9) | 1 | 56 |
| Resilience | The Generalised Anxiety Disorder (GAD-7) | 1 | 56 |

### Objective 2: Model development

#### Outcomes

All identified outcomes were reported as change for healthcare staff. Three papers reported a change in the therapeutic staff-service user relationship rather than the healthcare staff individually, listed under the category most relevant to reported outcomes. Outcomes reporting a change in staff-service user relationships describe a strengthened therapeutic alliance (55-57). Most papers identified helpful changes to healthcare staff and services (Table 9).

**Table 9.** Helpful outcomes identified in included studies, arranged by higher-level category and sub-category. Multiple papers from the same study were counted separately only if reporting different outcomes. Some outcomes were described ambiguously in their original papers and therefore included in, but not expanded on, in the table.

| Higher category | Outcomes | Study ID(s) |
| --- | --- | --- |
| Short-term emotional change for healthcare workers | Boosted confidence | 40, 41, 65 |
|  | Boosted morale | 38, 65 |
|  | Confirmation of doing good work | 16, 18, 25, 28, 33, 41, 42, 50, 58, 62 |
|  | Coping resource at work | 55 |
|  | Enthusiasm for the job | 54 |
|  | Experience of having a good day | 50, 60 |
|  | Feeling comfortable in their job | 65 |
|  | Feeling empowered | 7 |
|  | Feeling encouraged | 5, 11, 28, 42, 45, 54, 55 |
|  | Feeling engaged | 50 |
|  | Feeling fulfilled | 6, 39 |
|  | Feeling good | 38, 40, 41, 48, 51, 65 |
|  | Feeling happy | 10, 24, 29, 38, 39, 55 |
|  | Feeling honoured to serve their community | 33 |
|  | Feeling inspired | 1, 54 |
|  | Feeling positive about work | 51, 53, 63 |
|  | Feeling proud of work | 2, 6 |
|  | Feeling rewarded | 1, 6, 20, 24, 29, 40, 43, 44, 45, 54, 55, 59, 62 |
|  | Feeling successful | 10 |
|  | Feeling supported | 7 |
|  | Feeling valued | 2, 7, 36, 55, 58, 63 |
|  | Feelings of hope | 26 |
|  | Feelings of love for work | 30 |
|  | Feeling that the reciprocal respect between service user and healthcare worker is fulfilled | 52 |
|  | Increased individual energy at work | 30, 58 |
|  | Increased gratification | 22, 33, 48, 68 |
|  | Increased gratitude of healthcare workers | 6, 55 |
|  | Increased motivation at work | 2, 5, 6, 11, 21, 23, 24, 27, 28, 30, 32, 37, 38, 39, 42, 46, 47, 54, 55, 65, 67 |
|  | Increased personal satisfaction | 51, 58 |
|  | Increased psychological wellbeing | 6, 7, 36, 56, 58 |
|  | Increased sense of achievement | 45 |
|  | Greater self-reflection about practice | 6 |
|  | Source of strength/support during difficult times | 6, 28, 68 |
| Work-home interactional change for healthcare workers | Improved familial satisfaction for spouses of healthcare workers | 57 |
|  | Improved work-home relationship | 57 |
| Work-related change for healthcare workers | Created a positive work environment | 61 |
|  | Improved communication with service users | 32 |
|  | Improved team diagnostic and procedural performance | 49 |
|  | Increased commitment to work | 28, 31, 54, 55 |
|  | Increased connection to service users and families | 50, 68 |
|  | Increased intention to refer to a service being positively evaluated | 15 |
|  | Increased sense of doing meaningful work | 16, 24, 45, 50 |
|  | Increased staff retention | 3, 6, 8, 9, 14, 38, 63, 64 |
|  | Increased work-related activity | 4 |
|  | Increased work-related satisfaction | 1, 6, 9, 13, 17, 20, 23, 24, 28, 31, 32, 33, 34, 38, 43, 46, 48, 50, 51, 54, 55, 58, 59, 61, 62, 63 |
|  | Reduced burnout | 6, 19, 56 |
|  | Reduced perception that assigned tasks are avoidable or outside of job role responsibility | 37 |
|  | Strengthened therapeutic alliance | 35 |

Some papers identified undesirable changes (Table 10).

**Table 10.** Undesirable changes for healthcare staff identified in included studies.

| Change category | Sub-category | Study ID(s) |
| --- | --- | --- |
| Short-term emotional change for healthcare workers | Feeling embarrassed when being delivered feedback from tutors | 41 |
|  | Feelings of envy and stress when not rewarded with positive feedback | 55 |
|  | Feelings of guilt after accepting a gift | 35 |
|  | Feelings of tension and pressure to meet philanthropic service user expectations | 66 |

One change was identified which could be viewed as both helpful and undesirable. An altered responsiveness to grateful service users who give philanthropic gifts could be viewed as helpful as responding more quickly to those giving gifts may increase the likelihood of future donations (56). However, altered responsiveness may undermine the professional relationship between staff and service-users and result in a decreased responsiveness to those not giving gifts.

#### Mechanisms

A mechanism is a process by which positive feedback causes change. Mechanisms identified in included studies are in Table 11.

**Table 11.**
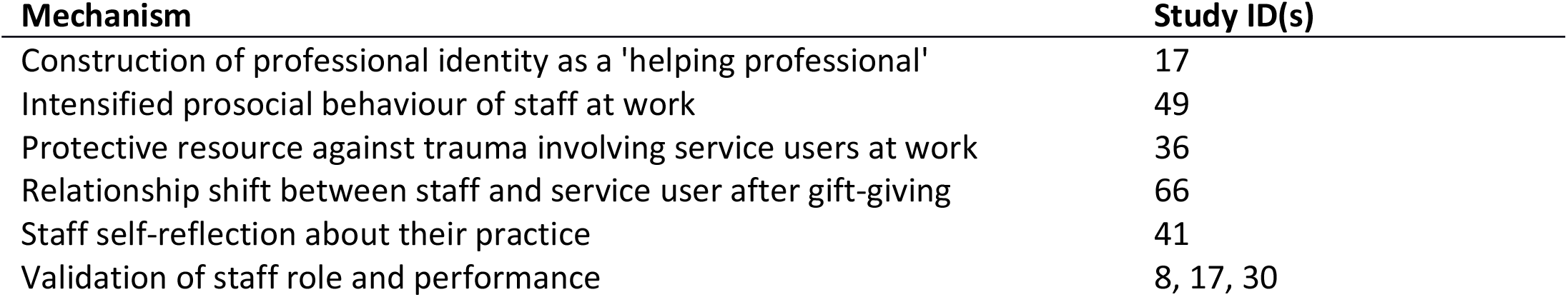
Mechanisms identified as cause of change in included studies.

#### Moderators, facilitators, and barriers

Factors were identified which can alter the degree of change following positive feedback. Some factors enhanced the effect of positive feedback (Table 12).

**Table 12.**
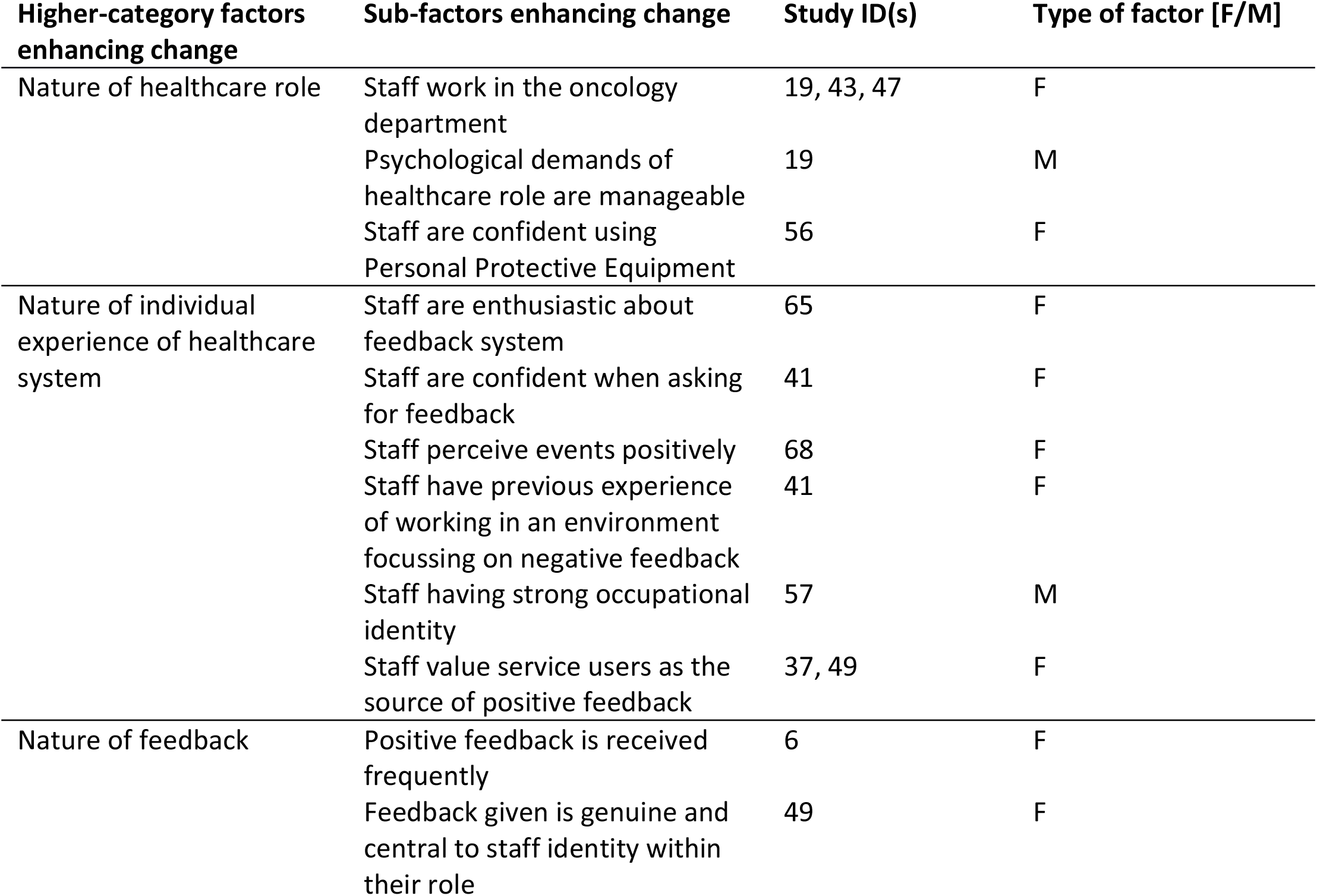
Factors enhancing the effect of positive feedback in included studies. Type of factor refers to facilitators [F], or moderators [M].

Some studies also identified barriers to change, where the effect of positive feedback was hindered (Table 13).

**Table 13.**
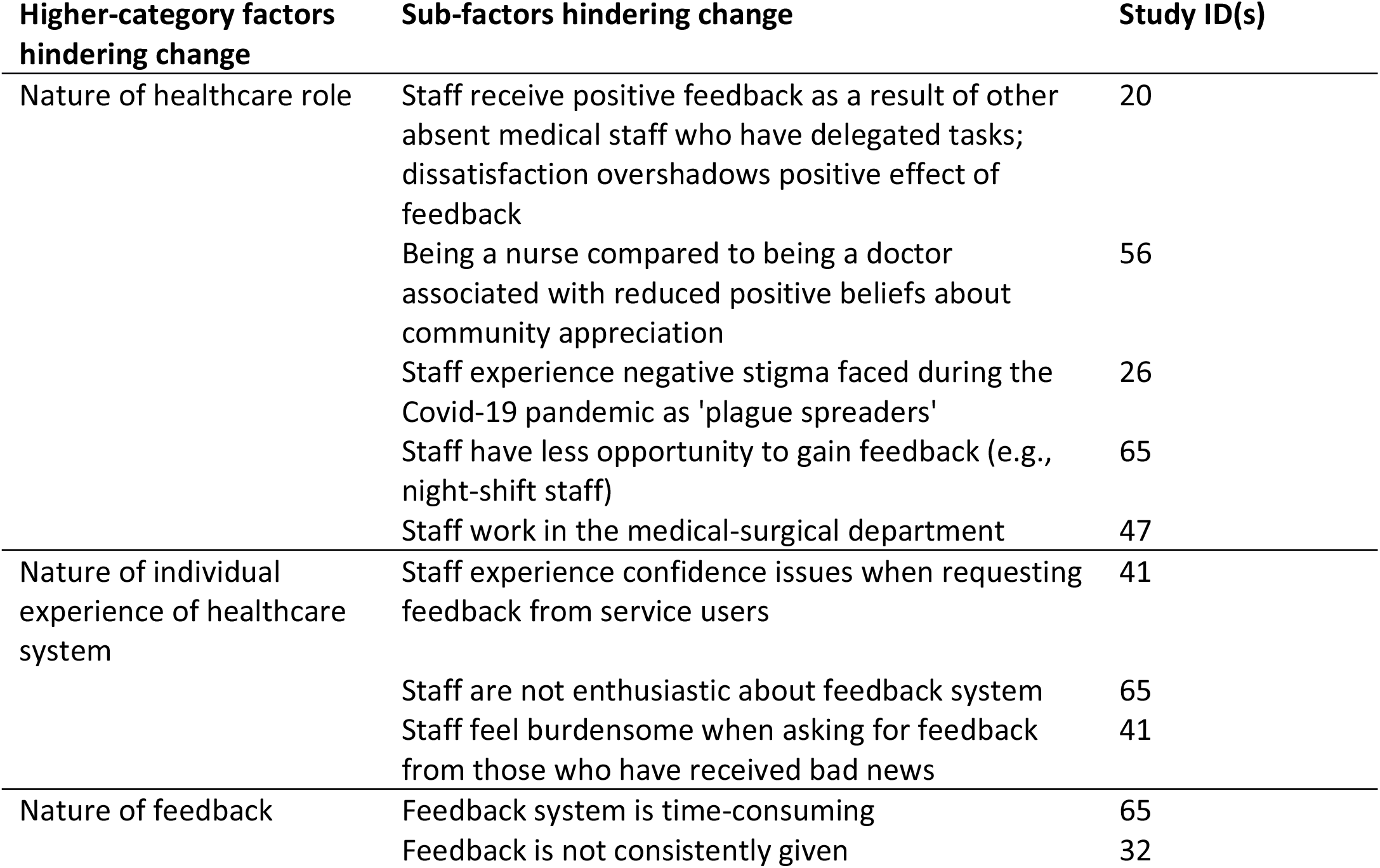
Factors hindering the effect of positive feedback in included studies.

| Higher-category factors hindering change | Sub-factors hindering change | Study ID(s) |
| --- | --- | --- |
| Nature of healthcare role | Staff receive positive feedback as a result of other absent medical staff who have delegated tasks; dissatisfaction overshadows positive effect of feedback | 20 |
|  | Being a nurse compared to being a doctor associated with reduced positive beliefs about community appreciation | 56 |
|  | Staff experience negative stigma faced during the Covid-19 pandemic as 'plague spreaders' | 26 |
|  | Staff have less opportunity to gain feedback (e.g., night-shift staff) | 65 |
|  | Staff work in the medical-surgical department | 47 |
| Nature of individual experience of healthcare system | Staff experience confidence issues when requesting feedback from service users | 41 |
|  | Staff are not enthusiastic about feedback system | 65 |
|  | Staff feel burdensome when asking for feedback from those who have received bad news | 41 |
| Nature of feedback | Feedback system is time-consuming | 65 |
|  | Feedback is not consistently given | 32 |

Three studies described working in oncology as enhancing the effects of positive feedback. One study described having increased intimacy and closeness with oncology service users, facilitating feelings of reward and satisfaction (58). Another described how working in oncology felt more worthwhile and like a gift, with service users expressing deep appreciation which is not seen in other wards. In comparison, working in a medical-surgical unit can feel robotic (59).

One study described how working in oncology had fewer psychological demands (60). The psychological demands of the healthcare role impacted the degree of change between service user gratitude and burnout. Emergency units were perceived to have higher psychological demands than oncology wards, due to work shifts, workloads, and the shorter, more superficial relationships with service users. For emergency nurses, personal accomplishment as a mediator of burnout diminished with increased psychological demands. In contrast, oncology nurses had higher perceptions of service user gratitude and higher personal accomplishment. The institutional context may influence the extent to which staff members are able to encounter and engage with positive feedback.

Occupational identity was also identified in another study as factor enhancing the effect of service user gratitude, with changes to energy within relationships, spousal family satisfaction, and relationship-based family performance (61). Receiving service user gratitude improved healthcare staff’s home environment, and this was amplified when staff strongly identified with their role.

In one study, appreciation reduced the relationship between intrinsic motivation (a type of motivation that is based on inherent pleasure or passion, rather than extrinsic rewards such as money or fame) and the perception of illegitimate tasks (62). Illegitimate tasks were unnecessary (tasks that could have been avoided with better organisation) or unreasonable (tasks that were not the responsibility of that staff member). Motivated staff perceived a higher number of unnecessary tasks being assigned to them, but appreciation from service users reduced this relationship.

#### Mediators

A mediator is a factor which is essential in the change process and must be in place for change to occur. In the study by Riskin et al (2019), team information sharing partially mediated the impact of gratitude (63). In a study by Tang et al (2021) energy within relationships mediated the effect of service user gratitude and spousal family satisfaction (95% CI = 0.007 to 0.042; *P* = .02) and relationship-based family role performance (95% CI = 0.010 to 0.054; *P* = .03) (61). Receiving gratitude from service users acts as an energy resource within relationships, which healthcare staff are then able to utilise in the family domain. As a result, increased relational energy led to increased familial satisfaction.

### Change model

Forms of change are summarised in a model presented in Fig 2, which extends a preliminary model from a prior review (37).

**Fig 2.**
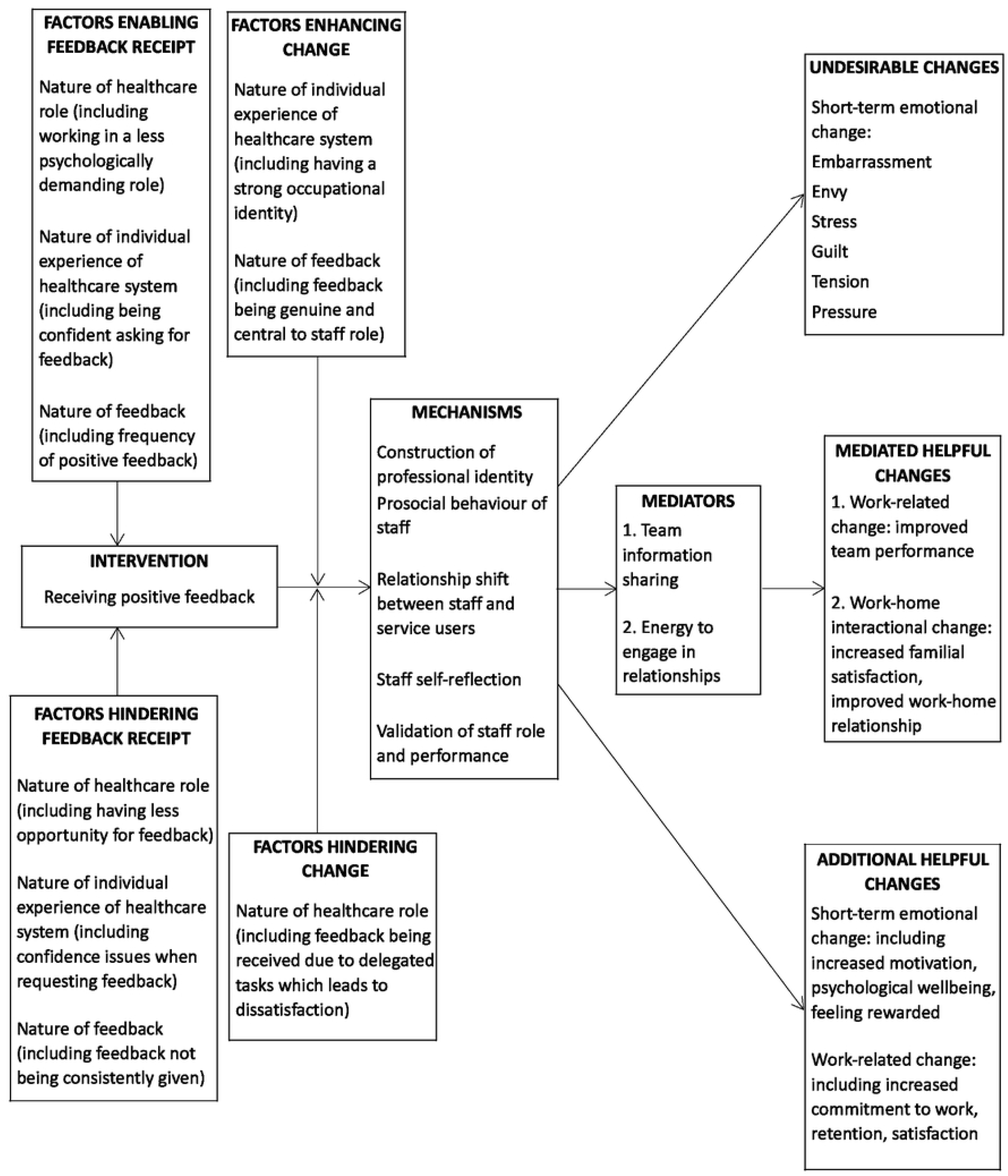
How positive feedback from service users, families, and communities can create change in healthcare settings conceptualised in a change model

Listed factors are not exhaustive.

### Subgroup analyses

#### Quality assessment

Only one study (reported on in two papers) did not meet the 60% threshold for quality assessment due to a lack of a clear research question (50, 64). Findings from this study were not consequential to the change model due to these being reinforced by other studies (50).

#### Studies conducted in a mostly public versus mostly private healthcare system

One difference between studies conducted in a mostly public healthcare system (UK) and mostly private healthcare systems (US) was the type of positive feedback provided. All UK studies described ambiguous types of positive feedback. While many US studies also described ambiguous feedback, two described material feedback in the form of cards, flowers, and gifts (56, 65). One undesirable change was identified in both UK and US studies. In the UK, research identified that students feel embarrassed when receiving positive feedback from feedback forms via tutors (66), whereas in the US, tension and pressure surrounding the service user-professional relationship was identified after gift-giving (56).

## Discussion

### Summary of findings

The review included a broad range of papers presenting evidence that change can be created in health services using positive patient feedback. The largest body of evidence relates to beneficial short-term emotional changes experienced by healthcare workers as the result of receiving feedback, such as feeling more hopeful and motivated, and to beneficial work-related change (such as increased retention and reduced burnout). Beneficial changes to the home environment were also documented. A small number of undesirable changes were identified. These included feeling embarrassed when receiving feedback, feeling envy and stress when not rewarded with positive feedback, and feeling guilt, tension, and pressure when accepting gifts. Tensions surrounding service user gift-giving may arise due to health professionals being restricted to only accepting ‘trivial’ gifts, which may create uncertainty in staff regarding boundaries due to vague definitions (67). The type of gift (such as those marking an occasion, inexpensive, or ‘over the top’) and recipient (such as individual staff or donation to the service) may influence staff reactions. Gifts which fail to align with ethical practice, such as ‘over the top’ displays of gratitude, may be more likely to produce undesirable change (68).

Importantly, only two intervention studies were identified (63, 65), and neither quantified effect in a real-world healthcare setting. This means that no evidence on the size of effect produced by positive feedback was available. This points to a substantial gap in knowledge which might be addressed by future research studies. A broad range of measures were used in quantitative studies, suggesting a lack of consensus in the research community on the most important constructs to consider, and how to assess them. Most work has been conducted within the last 10 years, which potentially relates to the widespread emergence of technological solutions to the collection and distribution of feedback, creating the potential for new forms of intervention.

The current review has identified factors which enhance or hinder the creation of change through feedback. Some of these factors relate directly to the nature of specific healthcare roles and professions. For example, change was enhanced if feedback recipients worked in roles which allow more meaningful interaction with service users, and hindered for feedback recipients working night shifts and hence potentially having less direct contact with patients. This suggests that positive feedback may not be an accurate measure for assessing quality of care as some staff are not given the opportunity to influence and receive feedback. It is unlikely that feedback will be equally received by staff across services due to their varying nature with the implementation of a single feedback system. Tailoring feedback systems to the settings and contexts in which staff work may be beneficial to ensure similar opportunities to receive feedback but understanding the fundamental differences between services is crucial when assessing quality improvement priorities.

### Relationship to prior work

The current review extends a previous systematised review which investigated how expressions of service user gratitude creates change in healthcare services (37). Due to the current review having a mostly broader focus, 68 papers were included compared to 26 papers in the previous review, and this has resulted in a broader range of short-term emotional benefits and undesirable impacts being identified.

In a scoping review investigating service user gratitude in healthcare, receiving gratitude was found to enhance healthcare worker wellbeing, act as a positive force against stress, increase motivation, increase reciprocated gratitude, and reduce burnout (35). Aparicio and colleagues identified 32 includable papers, only two of which were included in the current review (59, 60). Despite a lack of cross-over in included studies due to differences in inclusion criteria, the findings remain consistent. For instance, gratitude acting as a positive force against distress is also seen in the current review, categorised as increased psychological wellbeing and a protective force against trauma. The methodology of the current review builds on the scoping review by examining literature systematically, in line with MRC recommendations (38).

The benefits of positive feedback identified in this review may be particularly relevant for the occupational health of healthcare staff. For example, the number of nurses leaving the profession rose in 2021 by 25% (69), with increased workload leading to higher levels of burnout (70). Healthcare workers have been found to have high levels of intrinsic motivation, where motivation to perform well is a product of inner drives. This was particularly evident in permanent healthcare staff (71). Validation of having done good work may therefore be positively reinforced with positive feedback and be of greater value than for those who are extrinsically motivated by factors such as financial reward or promotion (72). Increased intrinsic motivation may boost affective commitment and lead to reduced turnover intention among healthcare staff (73). Similarly, finding intrinsic meaning in their work was helpful for healthcare workers in Japan to cope during the Covid pandemic (24)). Self-determination theory also suggests that intrinsic motivation can assist with the development of professional identity for healthcare staff (74).

Shifting healthcare staff attitudes surrounding service user feedback may be essential for implementing meaningful change which utilises positive feedback due to the belief that feedback is largely negative (13). The Lewin Change model describes three steps for creating change (75), starting with ‘unfreezing’ whereby a shift away from current beliefs is initiated through challenging defensiveness towards change and dismantling current views. This may be possible through exposure to positive feedback. The second stage is ‘movement’ which describes a change occurring, such as beneficial outcomes as a result of positive feedback. The third stage is ‘refreezing’ which describes a replacement of old views and processes with new ones, which begins to normalise the new methods of operating. For positive feedback in healthcare, this may reflect system-level change such as policy implementation.

However, this model may be limited to healthcare staff’s willingness to engage with positive feedback. The idea of a ‘learning organisation’ was introduced by Senge, who described a group of people continually working to enhance their capacities and create results that they want (76). A learning organisation describes one which is not operating as a machine, but rather a humanistic never-ending process of development and learning. Adapted for healthcare settings, learning organisations have five disciplines (77). ‘Open systems thinking’ describes services being viewed as a whole rather than isolated by disease, procedures, or structures, and aims to create interconnectedness beyond departmental boundaries. ‘Improving individual capabilities’ describes striving for excellence by improving personal proficiencies of staff. ‘Team learning’ describes learning as a collective rather than via single professionals. ‘Updating mental models’ describes updating the deeply held assumptions and generalisations held by individuals within the organisation and finding new ways of operating. Finally, ‘a cohesive vision’ describes empowering and enabling staff being counterbalanced by strategic direction and clear values to guide individual action to produce shared understanding. Healthcare systems have identified that being a ‘learning organisation’ encourages a culture celebrating innovation and success (77). Positive feedback may offer a means for learning organisations to create a cultural shift towards valuing positive service user experiences rather than focussing solely on negative incidents and risk reduction.

### Strengths and limitations

A strength of the review is that a broad range of publications databases was searched, including a database specific to computing publications and rarely used in systematic reviews, which is important when feedback is routinely collected through technological means. Compared to the prior narrower review, broader inclusion criteria have enabled the inclusion of papers describing changes to healthcare systems, enabling the identification of changes such as increased referral intentions following positive feedback from service users about a particular service (78). The addition of search terms such as ‘positive feedback’ and ‘positive evaluation’ have enabled new forms of change to be identified, such as non-clinical staff benefiting from positive feedback as well as those in clinical roles. A strength of the review is that inclusion criteria were carefully designed to exclude papers where there was ambiguity about the source of feedback or the direction of change, meaning that studies were excluded where causality was uncertain, such as in studies using correlation analyses (79). This has provided a solid foundation to develop a change model.

Another strength of the review is that it was inclusive of studies which were conducted in non-WEIRD (western, educated, industrialised, rich, and democratic) countries, despite being limited to papers published in English. For example, included studies reflected healthcare systems in eight African regions. Although emotional expressions differ across cultures (15), positive feedback was deemed helpful to healthcare organisational outcomes. Findings were robust across studies despite differing locations and healthcare systems, reinforcing the value of positive feedback.

A limitation of the review is that the definition of positive feedback is not straightforward. A subgroup analysis was planned for documents which identify change through expressions of healthcare service user gratitude specifically. Ambiguity in the distinction between positive feedback and gratitude definitions meant that the subgroup analysis could not be performed. Medical definitions of positive feedback describe the body being amplified from its normal state (80), but this review did not include positive physical or medical signals from service users. However, seeing a patient improve was described in some studies as a form of positive feedback

(81). Physiological markers may not reflect positive healthcare experiences and would not reflect quality of care given by palliative care teams. Further, service user gratitude was seen to create change for other service users (20), but this was excluded as it could not be considered a change for healthcare staff or systems.

Furthermore, positive feedback was defined as a response from healthcare service users, families or the community indicating concordance between desired and actual experiences regarding their care or treatment, delivered to healthcare staff or systems. However, the assumption was made that positive feedback was expressed with the intention of communicating this concordance between desired and actual care, but other contextual and motivating factors may have existed, such as feeling obligated to give positive responses when asked for feedback in person (82), service users attempting to influence their future care and treatment and prevent punitive treatment for negative feedback (20), and social norms surrounding expressions of thanks which may be expressed habitually (83).

### Implications of the review and change model

#### Implications for practice

Managers of health service units seeking to address problems such as staff burnout or low motivation should consider the integration of mechanisms for making positive feedback available to staff members and should seek to identify barriers to the use of positive feedback in their units. Health service managers in units already making use of positive feedback should examine whether particular staff groups are disadvantaged, for example if working in circumstances that make the provision of positive feedback more difficult, or increasing exposure of positive feedback to individuals from minority ethnic backgrounds who may be more likely to receive complaints (84). Policymakers should consider adopting policies that encourage the collection and distribution of positive feedback. Requirements of healthcare professional bodies to make use of feedback in reflective practice might be used to motivate change, though it is unclear whether this phenomenon extends beyond the UK. This may also exclude individuals whose roles do not require professional registration.

Integrating positive feedback from service users, families, or communities into standard clinical supervision rather than formal requirements may create an attitudinal shift away from revalidation scepticism to become an essential part of practice (28). Effective clinical supervision can prevent burnout (85), and positive feedback may enhance these benefits.

#### Implications for research

There is a need for the research community to conduct and replicate intervention studies of precisely designed feedback interventions, ideally in the form of randomised controlled trials designed to collect evidence on effectiveness and cost-effectiveness. The research community should seek to reach consensus on the most important measures to be assessed in trials, to enable meta-analyses pooling findings from multiple trials to be conducted. Future research may investigate the effects of positive feedback depending on healthcare role, comparing those who have consistent access to feedback (such as oncology staff) (59), to those who feel overlooked and undervalued (such as healthcare assistants) (86). Future research may investigate the effects of positive feedback at multiple levels of the organisation, such as individual impacts like resilience, and organisational culture and system-level change, and whether the effect of positive feedback changes depending on individual or team receipt.

The research community should also aim to investigate the influence of feedback content and form in eliciting change and whether content has practical utility. Examples include whether content of feedback is meaningful to staff, and if relationships with service users are more significant than numerical indicators of satisfaction. Feedback with specific utility, such as an appointment being ‘on time’, may also produce differing effects to interpersonal emotional connections. This may assist with the development of a typology to characterise feedback and assist with understanding whether positive feedback should be used and delivered universally.

Research may also benefit from being co-designed with healthcare workers with practical knowledge to enhance the functional integration of findings into clinical practice.

## Conclusions

Positive feedback from service users, families, and the community can lead to meaningful change. Change is largely positive, with emotional, familial, and work-related change being recognised. However, some undesirable changes were identified in relation to healthcare staff emotions. Overall, positive feedback provides the opportunity to create meaningful change but requires further interventional investigation.

## Data Availability

All relevant data are within the manuscript and its Supporting Information files.

## Acknowledgements

For the purposes of open access, the authors have applied a CC BY public copyright license to any author accepted manuscript version arising from this submission

## Supporting information captions

S1 File. Amendments to search strategy for CINAHL, ASSIA and the ACM Digital Library

S2 File. PRISMA checklist

S3 Table. Full data abstraction table

